# Beyond the Gene in Genetics: How Isoform-Resolved Analysis Empowers the Study of Both Common and Rare Genetic Variation

**DOI:** 10.1101/2025.03.31.25324931

**Authors:** Jeppe Malthe Mikkelsen, Dimitrios S. Kanakoglou, Chunxu Han, Peter Wad Sackett, Simone Tiberi, Kristoffer Vitting-Seerup

**Author notes:** Authors contributed equally and can freely change their order within their CVs (Jeppe Malthe Mikkelsen, Dimitrios S. Kanakoglou).

## Abstract

**Background:** Genetics is rapidly deepening our understanding of human health and disease by investigating both common and rare genetic variants and their influence on gene expression[1,2]. Alternative splicing is a molecular mechanism enabling a single gene to produce multiple protein isoforms with distinct biological functions[3]. However, while protein isoforms are ubiquitous in humans, we here show that they are rarely considered when investigating common or rare genetic variants.

**Methods:** To address this gap, we systematically assessed the impact of isoform-level analysis across 83 Quantitative Trait Loci (QTL) datasets and 76 million human genetic variants.

**Results:** Using strict colocalization, we find that ∼46% of genes associated with human phenotypes via Genome-Wide Association Studies (GWAS) are associated through isoform-level expression changes. Similarly, we find that 72% of rare pathogenic variants are interpreted differently depending on the isoform expressed in relevant tissues and cell types.

**Conclusions:** These findings establish isoform-level analysis as an essential approach for genetic research. Furthermore, they underscore the need to (re)analyze genetic data at isoform resolution, thereby unlocking deeper insights into human biology and expanding clinical applications.

## Background

The efforts to understand human genetic variants are often divided into analysis of common and rare variants based on how frequent the variant is in the general population[4]. Common variants of interest are primarily identified through Genome-Wide Association Studies (GWAS). Most GWAS-identified variants are, however, located in non-coding regions and thereby lack functional interpretation. Therefore, through colocalization analysis, GWAS results are often integrated with Quantitative Trait Loci (QTL) data, thereby linking GWAS variants to specific genes[5]. In contrast, rare variants are typically identified through genome sequence of people with inherited disorders, specific cancer types, or rare diseases with a focus on identifying pathogenic variants within genes[6]. Thus, both research fields have provided extensive and fundamental insights into the molecular mechanisms underpinning the human condition by investigating the genetic effect of the genes associated with a variant[7,8].

Most human protein-coding genes give rise to multiple distinct protein isoforms[9,10]. Among these genes, 93% have been shown to significantly alter their isoform composition in context- and condition-specific ways[11]. This also means that isoforms are involved in almost all cellular processes, with apoptosis among the best described. In the apoptotic signaling cascade, most central proteins utilize anti-apoptotic isoforms under normal circumstances but switch to pro-apoptotic isoforms when apoptosis is activated[12]. As described above, investigating genetic variants relies heavily on interpreting the function of genes associated with specific variants. It is, therefore, striking that isoforms are only considered in a minority of studies considering GWAS, QTL, and rare variants. Specifically, our literature review finds that less than 20% of recent genetics studies consider isoforms regardless of whether they analyze common or rare variance (Figure 1A, Supplementary Table 1). This is especially important since examples where isoforms have different biological functions and can change the interpretation of a variant are well-known[13–16]. Although recent landmark studies have demonstrated that common and rare genetic variants frequently affect splicing[17,18], it is still unclear how critical it is to consider isoforms when analyzing common and rare genetic variants. To systematically quantify this, we here used an isoform-centric analysis approach to re-analyze most publicly available RNA-seq QTL, GWAS studies[19,20], as well as all possible missense and pathogenic variants in the human genome[21–23].

**Figure 1:**
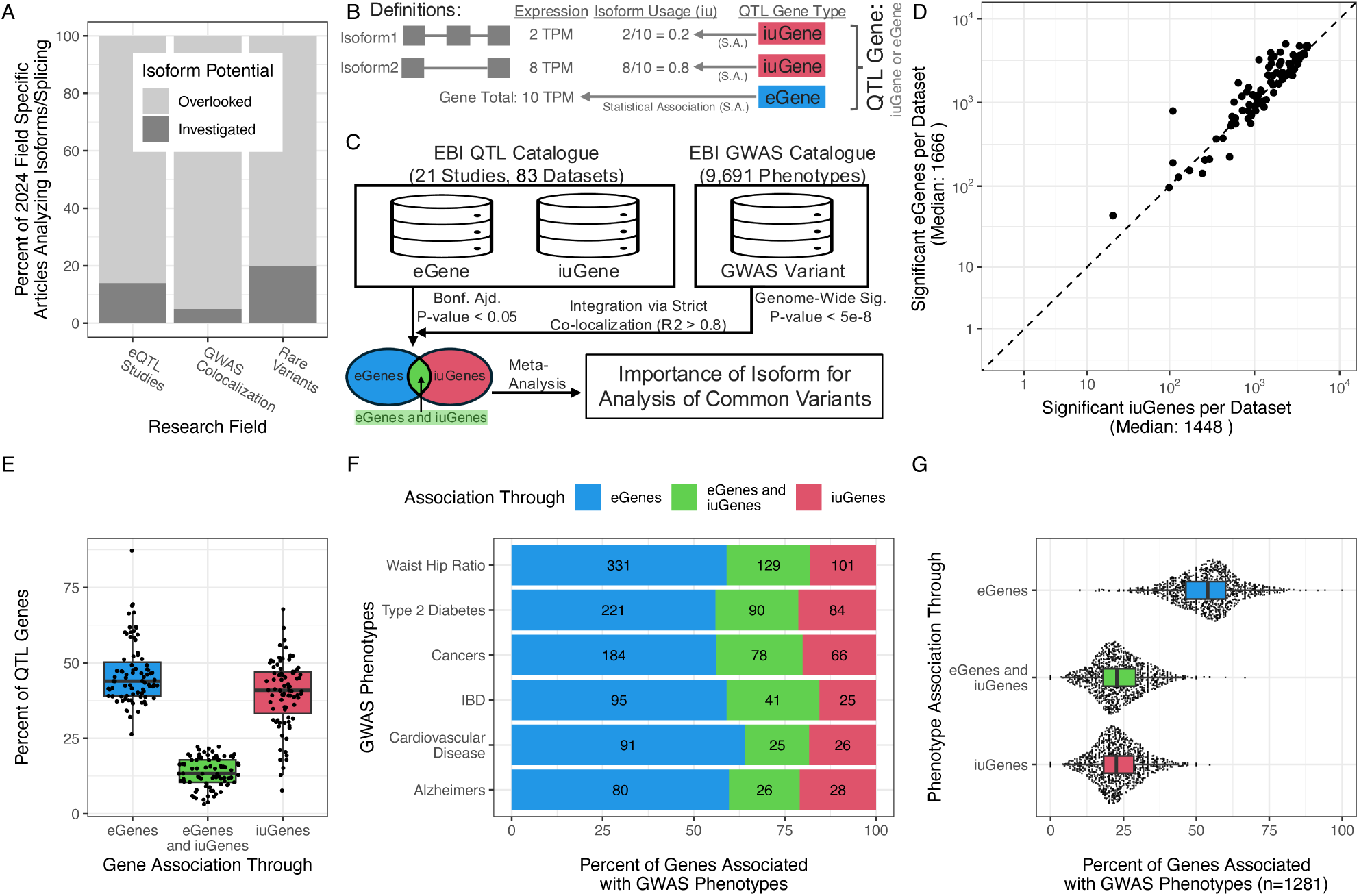
RNA-based isoform analysis empowers QTL and GWAS studies. **A)** Literature review of how frequently isoforms are considered (y-axis) for various research fields or analysis approaches (x-axis) (See Supplementary Table 1). **B)** Definition of iuGenes, eGenes, and QTL gene. The color indicates color usage in downstream analyses. **C)** Schematic overview of data integration for gene-level analysis and cutoffs used. Color as in B. **D)** Per dataset comparison of the number of eGenes and iuGenes. **E)** Per dataset, the percent of significant QTL genes (y-axis) being eGenes, iuGenes, and both (x-axis and color). **F)** For selected GWAS phenotypes (y-axis), the percent of genes associated with the phenotype through variant colocalization (x-axis) with color indicating the QTL type gives rise to the association. Superimposed numbers show the underlying number of genes. **G)** generalization of F) to 1,281 GWAS phenotypes, each having at least 10 colocalized genes.

## Methods

### Literature review

The literature was retrieved from PubMed using the RISmed R package (v2.3.0). For clinical variants, the literature review was done using the following search criteria: (i) publications between January 2022 and December 2023; (ii) article type restricted to “Journal Article”; (iii) studies on human; and (iv) titles or abstracts containing the terms “rare variants” and either “RNA-seq” or “RNA sequencing.” The collected articles were randomly ordered and reviewed until 50 studies that met the inclusion criteria were identified. Specifically, the inclusion criteria were (i) a study of humans, (ii) the full-text article was available to us for review, (iii) verifying it was a QTL study, and (iv) verifying the study used RNA-seq data, thereby having the potential for isoform analysis. Each of these 50 articles was further examined to determine if they included splicing analysis. Such analysis typically encompasses quantifying isoform or transcript expression, identifying alternative splicing events or changes, and other analyses at the isoform or transcript level rather than at the gene level. The fraction of studies incorporating splicing analysis was calculated by dividing the number of articles addressing splicing by the total number of reviewed articles (n=50). For eQTL and co-localization studies, the same approach was used searching for respectively “Quantitative Trait Loci” and “genetic colocalis(z)ation” or “colocalis(z)ation analysis”. For the co-localization, we furthermore specified the query to not include “fluorescence,” we reviewed 100 co-localization studies to ensure the estimate was more precise (as co-localization had fewer articles analyzing isoforms).

### QTL Analysis

#### Data used

Permutation *P*-value files (*.permuted.tsv.gz) were retrieved from the EBI QTL Catalogue[19] for all studies listed in Supplementary Table 2, resulting in two separate datasets for each study (one for gene-level expression associations and one for transcript-usage-level associations). As detailed in Kerimov et al., [19] gene-expression was obtained by aligning RNA-seq reads to the genome and counting reads overlapping with featureCounts, while transcript usage was derived from Salmon quantification by dividing isoform TPM values with total TPM from all transcripts of the same gene. Both pipelines used Gencode v39 as reference. All associations between variants and RNA-seq data were calculated using QTLtools, considering only pairs within 1 MB of each other and correcting for both genetic and phenotypic data-specific effects, as detailed elsewhere^1^.

#### Ancestry analysis

To confirm that all included datasets are of predominantly European ancestry, we for each dataset 1) computed the Pearson correlation between the observed Minor Allele Frequencies (MAF) of variants significant in at least one dataset (n = 87,189) and with available rsID mappings, and the 1000 Genomes Phase 3 reference superpopulation MAFs (obtained from Ensembl’s REST API) from 130,192 rsIDs (3.8% with missing population assignments were excluded), 2) and validated against an independent genotype-based PCA ancestry classification from the eQTL Catalogue (Pearson correlation: 0.875, *P*-value: 5e-9, 95% CI: [0.738, 0.943], R² = 0.765, 23/26 classification agreement; Supplementary Figure 1C.1, Supplementary Table 3). Reference allele frequencies from Ensembl’s REST API were converted to minor allele frequencies using pmin(AF, 1−AF) before computing correlations; QTL-side MAF values were verified to be ≤0.5 across all 109 analyses. MAF-based correlations served as a QC sensitivity check; exclusion decisions were based on donor-level PCA ancestry (eQTL Catalogue genotype-based PCA EUR% < 60%). Five parent studies were excluded, leaving 83 analyses across 20 retained parent studies, comprising 5,467 genotyped donors (92.0% European).

#### Analysis of significance

The 83 included QTL analyses span 75 distinct tissue and cell type contexts. This count was derived by extracting the tissue/cell type label from each analysis identifier (format: {study}_ge_{tissue_context}) and counting unique labels across all included analyses. Some tissues appear in more than one study (e.g., LCL in four studies: GENCORD, GEUVADIS, GTEx, and TwinsUK; blood in three studies: GTEx, Lepik_2017, and TwinsUK), which is why 83 analyses correspond to 75 unique tissue/cell type contexts. All 83 analyses use RNA-seq gene expression quantification (quant_method = ge) from the eQTL Catalogue; microarray-based studies (i.e., CEDAR, Kasela_2017, Fairfax_2014) were not included as they cannot be used for isoform-level analysis. For the datasets with transcript usage association, we focused on multi-isoform genes by removing all entries from genes annotated with only one transcript. The *P*-beta values for all entries were adjusted for multiple testing using the Bonferroni correction. A corrected *P*-value of 0.05 was used to identify genes with significant QTL association. eGenes and iuGenes were defined as genes with a significant association to an eQTL or transcript-usage QTL (in this article, referred to as isoform-usage QTL, aka iuQTL). Significant associations are available as Supplementary Data.

### Gene and variant overlap analysis

#### Gene-level overlap

To quantify the overlap between genes with significant eQTL and iuQTL associations, we used two complementary data sources: permutation-based significant gene lists (eGenes and iuGenes per condition, as described above) and purity-filtered credible sets downloaded from the EBI QTL Catalogue for all 83 retained conditions. For gene-level overlap, we paired each gene-expression (ge) condition with its corresponding transcript-usage (tx/txrev) condition by matching condition identifiers, yielding 83 paired datasets. For each paired condition, we computed the intersection and union of significant eGenes and iuGenes, and summarised the overlap as the percentage of genes in the union detected in both analyses, yielding a median overlap of 13.3% across all 83 conditions (Supplementary Table 7). Transcript-level molecular trait identifiers (ENST) were mapped to gene identifiers (ENSG) using the GENCODE v39 annotation.

#### Variant-level overlap

For variant-level overlap, we focused on the 42,025 gene-condition pairs co-occurring in both eGene and iuGene lists across all 83 conditions. For each such pair, we extracted all variants present in the ge and tx/txrev purity-filtered credible sets and determined whether at least one variant was shared. Gene-condition pairs with one or more shared credible set variants were classified as co-regulated (79.8%); pairs with no shared variants were classified as independently regulated (20.2%).

#### Distance analysis

For the distance analysis, we assigned each condition-gene observation to one of four categories based on significant gene lists and purity-filtered credible sets: eQTL only, iuQTL only, shared same variant, or shared different variants. For the variant-level analysis, we used condition-gene-variant observations from the relevant credible sets and calculated the distance from each credible-set variant to the nearest boundary of the associated gene body, assigning a distance of zero to variants located within the gene body. For the interval-level analysis, each credible-set genomic span was defined by the minimum and maximum variant positions in that credible set, and the distance from this interval to the associated gene body was set to zero when the interval overlapped the gene body. Distances were plotted as distance + 1 on a log10 scale so that gene-body and overlapping observations were retained in the visualization.

### Co-localization analysis

The “00.common.all” VCF-file from <u>NCBI</u> was used to identify rsIDs corresponding to the variant SNPs associated with significant e/iuQTLs as defined above. Specifically, this was done by looking up entries with matching genomic positions and mutations. Significant e/iuQTLs where no corresponding rsID could be found, and e/iuQTLs with the associated SNP located on a sex chromosome were omitted from the co-localization analysis (constituting ∼16-17% for both eQTLs and iuQTLs). To conduct the co-localization analysis we used the LDlinkR R package[24]. Specifically: the extracted rsIDs were given as input to LDlinkR::LDtrait(snps = <LIST_OF_RSIDS>, pop = “EUR,” genome_build = “grch38”, token = <ENTER_TOKEN>) in batches of 50 variants, using a token granted VIP access by the LDlink team to allow for parallel requests.

### Analysis of GWAS phenotypes

The result of the co-localization analysis was the association between variants associated with significant QTLs in our study and all phenotypes in the NHGRI-EBI Catalog of human genome-wide association studies (https://www.ebi.ac.uk/gwas/)[20]. We filtered GWAS associations to be of genome-wide significance (*P*-value < 5e-8) and with a stringent association between the GWAS variant and the variant found in our study (R2 > 0.8, a commonly used threshold for defining highly correlated variants likely to reflect the same underlying association signal[25,26]). The result was a database of stringent associations between genome-wide significant GWAS phenotypes and significant eGenes and iuGenes (available as Supplementary Data). Multiple phenotypes were concatenated for our analysis to better reflect the selected diseases shown in Figure 1F, as indicated by the table below. Figure 1G represents individual GWAS phenotypes, meaning no concatenation was performed.

For both Figure 1G and 1F, we, for each trait (or a combination thereof), counted the number of genes only associated with eGenes, the number of genes only associated via iuGenes, and the number of genes associated via both eGenes and iuGenes. To ensure the percentages reported were trustworthy, only phenotypes with at least 10 genes associated were considered for the final analysis. To ensure the result generalized to both disease and non-disease phenotypes, we manually annotated the 649 traits with most genes associated with them as “disease” (e.g., “type 2 diabetes”), “non-disease” (e.g., “height”), or “unknown” (e.g., “household income mtag”) (Supplementary Table 4).

| Figure 1F Phenotype | GWAS Phenotypes concatenated |
| --- | --- |
| Type 2 Diabetes | 'type 2 diabetes adjusted for bmi',<br>'type 2 diabetes' |
| Waist Hip Ratio | 'waisttohip ratio adjusted for bmi',<br>'waisthip ratio',<br>'waisthip index' |
| Cancers | 'prostate cancer',<br>'breast cancer', |
|  | 'colorectal cancer',<br>'cancer pleiotropy',<br>'nonmelanoma skin cancer',<br>'colorectal cancer or advanced adenoma',<br>'lung cancer',<br>'gastric cancer',<br>'breast cancer or lung cancer pleiotropy',<br>'cancer',<br>'skin cancer',<br>'epithelial ovarian cancer',<br>'cervical cancer',<br>'endometrial cancer',<br>'noncardia gastric cancer',<br>'pancreatic cancer',<br>'bladder cancer' |
| IBD | 'crohns diseas',<br>'inflammatory bowel disease',<br>'ulcerative colitis' |
| Cardiovascular Disease | 'cardiovascular risk factors',<br>'cardiovascular disease risk factors',<br>'cardiovascular disease' |
| Alzheimer's Disease | 'family history of alzheimers disease',<br>'alzheimers disease late onset',<br>'lateonset alzheimers disease',<br>'alzheimers disease or family history of alzheimers disease',<br>'alzheimers disease polygenic risk score upper quantile vs lower quantile',<br>'alzheimers disease' |

### QTL distances from SNP to gene

The distance from the SNP to the associated gene was calculated for each significant e/iuQTL. If the SNP was located upstream of the gene, the distance to the gene start was used; if downstream, the distance to the gene end was used. SNPs located within the gene body were assigned a distance of 0. For annotations of genomic positions, GENCODE[10] v39 genes were used by filtering for “gene” in the “type” column. Significant QTLs with no available distance information were excluded (less than 0.5% of all significant QTLs). Variants more than 100 nucleotides away from the gene (upstream or downstream) are defined as distal. To incorporate GWAS results into the length analysis, variants associated with genes that colocalized with any GWAS phenotype (as described above) were annotated as having a GWAS association.

### Analysis of major isoforms in proteomics

#### Datasets used

Isoforms can now be quantified from proteomics data when we know the abundance of the individual peptides[27]. To avoid the time and resource-intensive process of reprocessing raw proteomics datasets, we searched PRIDE (ebi.ac.uk/pride) for peptide files that had already been processed. Specifically, we searched for human datasets with the keyword “peptides.txt,” the peptide file produced by MaxQuant[28]. These peptide.txt files were downloaded from EBI’s FTP site for PRIDE[29] (ftp.pride.ebi.ac.uk). Each of these 252 files represents a dataset where intensity columns indicate samples. For each file, we extracted the peptide sequence, the Posterior Error Probability (PEP) value, all columns containing peptide abundance (The peptide intensity columns) and, when available, the number of missed cleavages, information about whether a peptide was reverse or a potential contaminant. Only datasets with at least 6 samples were considered. For each dataset, we first removed reversed peptides, peptides marked as contaminants, and peptides with >1 missed cleavage. Next, we corrected the pep values for multiple testing using FDR and only kept peptides with FDR < 0.01, as that is the standard within proteomics[30]. The remaining peptide sequences were mapped against unique GENCODE[10] v44 protein isoforms using the AhoCorasickTrie R package via the AhoCorasickSearch function using peptides as keywords, the isoforms as text, and specifying alphabet = “aminoacid”. Peptides that did not map to these isoforms were removed. Only the 56 datasets with at least 20,000 peptides passing all these filters were used in this study.

#### Proteomics quantification

Each sample in each of the 56 datasets (1,878 samples, Supplementary Table 5) was quantified individually using the R package IsoBayes[27]. Specifically, the peptide abundances for each sample were prepared with the load_data() function specifying input_type = “other” and abundance_type = ‘intensities’ along with the isoform annotation for each peptide. Afterward, the inference() function was run, supplying the Gencode v44 isoform:gene relations to the map_iso_gene argument. For each Isobayes result, the Isoform quantified, the Prob_present (probability present), the Abundance, and the Pi_norm (fraction of abundance originating from each isoform) were extracted.

#### Counting major isoforms

To ensure we only counted reliably detected isoforms, we, for each sample, extracted isoforms with Prob_present >= 0.95 and Pi_norm > 0.1 from genes annotated with multiple protein-coding isoforms in Gencode v44[10]. Next, for each gene in each sample, we extracted the most abundant isoform (aka Dominant, defined by having the largest Pi_norm value), randomly choosing an isoform if multiple isoforms shared the largest Pi_norm value. To obtain the results observed in min X samples, we subset the dataset to isoforms, which are the major in min X samples. Then, we count how many genes had multiple isoforms passing this threshold for each X considered. This analysis was performed both after pooling all proteomics samples across datasets (Figure 2A) and separately within individual studies to assess dataset-level variation (Supplementary Figure 2A.6).

**Figure 2:**
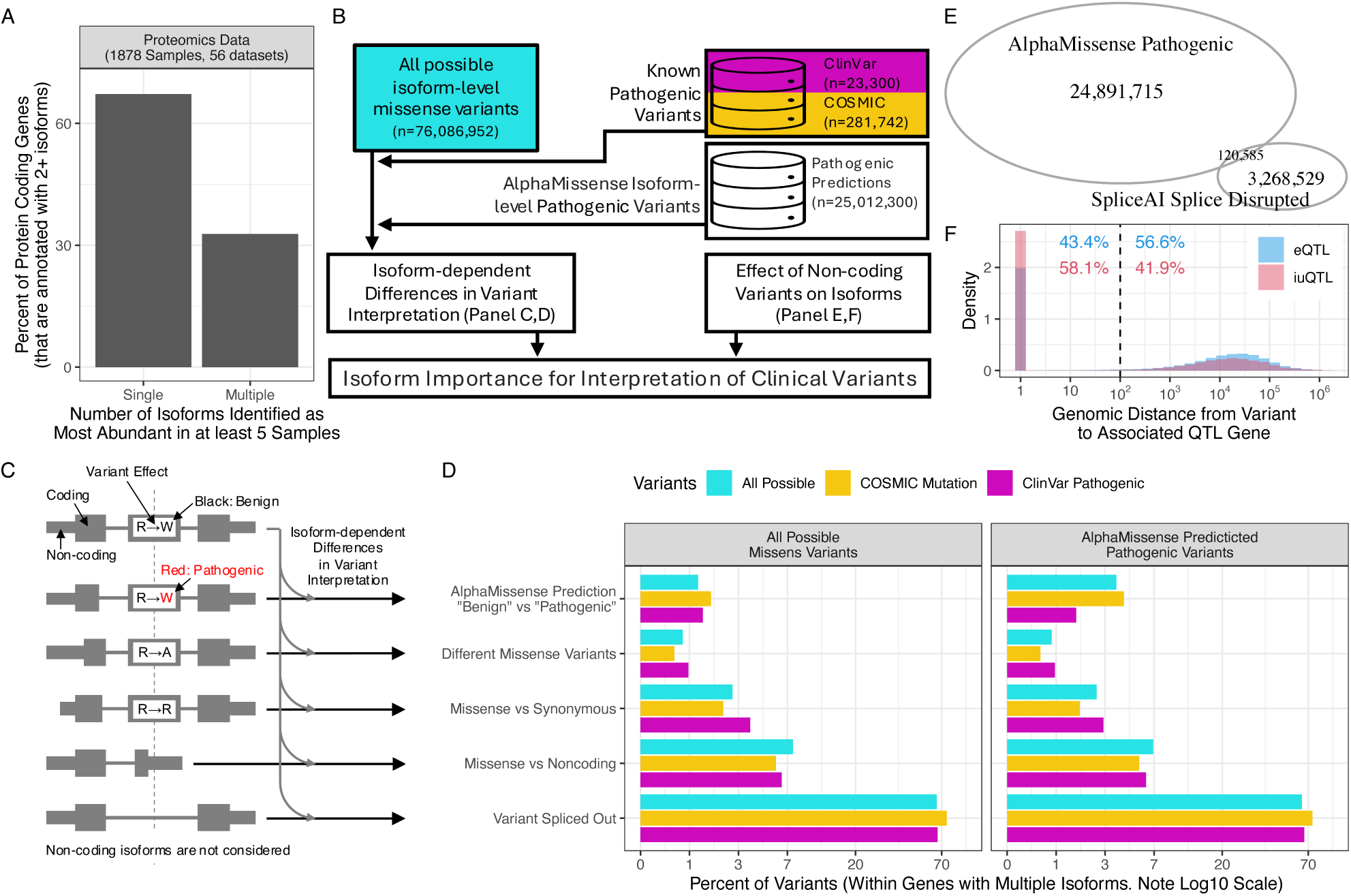
Importance of Protein Isoforms for Interpreting Rare Variants. **A)** The percent of detected multi-isoform protein-coding genes (y-axis) that had either one or multiple major (aka most abundant) isoforms identified in at least 5 samples. **B)** Schematic overview of data integration for rare variant analysis and colors used throughout the figure. **C)** Artificial schematic representation of how different isoforms can lead a missense variant (top isoform) to be interpreted differently when compared to the variant effect in isoforms (bottom 5 isoforms) with arrows indicating the corresponding analysis in D. **D)** The frequency of isoform-dependent differences in variant interpretations (x-axis) for each of the different categories considered (y-axis). Subplots and color jointly indicate the subset of variants considered. **E)** Overlap of pathogenic variants predicted by AlphaMissense and splice-disrupting variants predicted by SpliceAI. **F)** For each unique QTL gene, the genomic distance between the variant and gene. The color indicates QTL type.

Analysis of single corresponding proteomics dataset.

To assess the effect of the number of samples had on the observed number of genes with multiple major isoforms, we redid the analysis described above, but this time, we used 1,878 randomly selected samples from a recent database of Hela control proteomics samples[31].

#### Analysis of FDR Noise

The major concern for sub-gene level analysis of proteomics data is that each peptide, due to how MS/MS spectra are mapped to peptides, has a small chance of being wrongly assigned[32]. The cutoff almost always used allows a 1% False Discovery Rate (FDR) for assigning a spectrum to a specific peptide. In other words, 1% of peptides are probably wrongly assigned and most likely originate from another gene. Here, we term this problem “FDR noise” and assessed the impact of this on our proteomics analysis by adding 1% extra FDR noise and analyzing the effect. Specifically, for each dataset, we extracted the peptide abundance matrix and its associated peptide:isoform mapping data. We then randomly selected 1% of the peptides, and for each, we replaced the peptide:isoform mapping with another randomly chosen peptide:isoform mapping. In other words, we reassigned 1% of peptides to another isoform (and thereby also most likely to another gene). We then proceeded and quantified the data with added FDR noise (as described above) and counted the number of genes with multiple major isoforms (as described above). To ensure the results were not affected by outliers in the random sampling, this process was repeated 20 times, and the median number of genes with multiple major isoforms was extracted. Because of the computational intensity of this approach, this analysis was only done for a subset of the proteomics dataset (486 samples, 22 datasets, Supplementary Table 5). To compare the data with added noise with the actually observed data, we then re-counted the number of genes with multiple major isoforms (in the following termed signal) in the corresponding subset of the data. By subtracting the observed signal from the signal with added noise, we get the number of genes identified because of peptide FDR Noise. From the observed signal, we then define this number of genes as FDR Noise and the rest as a True signal.

### Analysis of major isoforms in RNA-seq data

To quantify dominant isoforms in human RNA-seq data, we utilized the ARCHS4[33] database which provide isoform expression as estimated by Kallisto[34] using annotation from Ensembl 107. From ARCHS4 we extracted 10,291 RNA-seq studies from the (version 2.1_beta) (Supplementary Table 6), and for each study extracted the median isoform expression only considering isoforms annotated transcript_biotype == ‘protein_coding’ in Ensembl 107 (the annotation quantified in ARCHS4). To ensure we only counted reliably detected isoforms, we, for each study, only considered isoforms with a median TPM expression > 1. Next, for each gene in each study, we extracted the most abundant isoform (aka Dominant, defined by having the largest median TPM value). To obtain the results observed in min X studies, we subset the dataset to isoforms, which are the major in min X studies. Then, we count how many genes had multiple isoforms, passing this threshold for each X considered.

To overlap with “reference” transcript databases, we downloaded APPRIS annotations from https://appris.bioinfo.cnio.es/#/downloads and MANE Select annotations from https://ftp.ncbi.nlm.nih.gov/refseq/MANE/MANE_human/release_1.5/ on 24th February 2026. For APPRIS, we extracted “PRINCIPAL:1” from the “APPRIS Annotation” column and for MANE, we extracted “MANE_status == ‘MANE Select’”. All analyses were subset to the 13,035 protein-coding genes shared between all 3 datasets. For each gene, we counted how many studies had transcripts not defined as “reference” by APPRIS or MANE Select as the most expressed (major) isoform in at least 3 samples.

### Analysis of isoform-specific missense variants

#### Obtaining AlphaMissense data

AlphaMissense[21] predictions from all possible transcript-level human missense mutations was downloaded from https://zenodo.org/record/8208688 by downloading the following two files: “AlphaMissense_hg38.tsv.gz” (isoforms termed “canonical” by the AlphaMissense authors) and “AlphaMissense_isoforms_hg38.tsv.gz” (isoforms termed non-canonical). The files were combined, removing 119 redundant isoforms from the non-canonical file. To combine with other datasets, we created the “var_id” which combines the chromosome number, genomic position, the reference, and the alternate. Please note that “var_id” is not transcript-specific but will be shared by all transcripts where the same missense variant was found.

#### Filtering AlphaMissense data

The combined AlphaMissense[21] data was filtered as follows: 1) Subsetted to transcript_ids also found in Gencode v32[10] (removed 470 transcripts), which the AlphaMissense data was created from according to the methods section. 2) Filtered for genes and transcripts classified as “protein_coding” in the Gencode v32 annotation (via the biotype columns) (This mainly, but not exclusively, removed transcript annotated as “nonsense_mediated_decay”).

#### Augmenting AlphaMissense data

In the following, we only considered Gencode v32[10] transcripts where at least one missense mutation passed all filtering described above (meaning we only consider isoforms observed in the AlphaMissense data). For each missense mutation in AlphaMissense data, we counted how many Gencode v32 transcripts had a genomic overlap (aka. exonic overlap, termed N_t-exon_) as well as how many transcripts had an overlap when only considering the annotated coding region (aka. CDS regions, termed N_t-coding_). Overlaps were counted via the GenomicRanges R package[35] by converting chromosome and genomic position (used for both start and stop) to Grange-objects and then using the countOverlaps() function. The number of non-coding transcripts overlapping a given missense variant was then defined as N_t-noncoding_ = N_t-exon_ - N_t-coding_. Next, we, for each gene, annotated how many Gencode v32 transcripts were observed at least once in the filtered AlphaMissense data (termed N_t-all_). These numbers allowed us to calculate two additional metrics: 1) The number of transcripts with overlapping coding regions without any missense variant, meaning it is a synonymous variant in that transcript: N_t-synonyms_ = N_t-all_ - N_t-coding_. 2) The number of transcripts where the variant was missing, meaning spliced out, part of an alternative transcription start site, or part of an alternative transcription termination site. We termed this number N_t-splice_ and calculated it as N_t-splice_= N_t-all_ - N_t-exon_. We then defined the “Pathogenic Variants” subset of the dataset as “var_id” where the variant from at least one transcript was predicted by AlphaMissense to be “likely_pathogenic”.

#### Integrating AlphaMissense and COSMIC data

COSMIC[23] data was downloaded from https://cancer.sanger.ac.uk/cosmic/. Specifically, the Cosmic MutantCensus version 98 for GRCh38 was downloaded on October 2nd, 2023. To integrate with the other datasets, we also created the “var_id” from the COSMIC data (see definition above). AlphaMissense predictions were then annotated as being in the COSMIC database by simply annotating if the constructed “var_id” was in both datasets.

#### Integrating AlphaMissense and ClinVar data

ClinVar[22] data was downloaded from NIH (https://ftp.ncbi.nlm.nih.gov/pub/clinvar/tab_delimited/). Specifically, “variant_summary.txt.gz” was downloaded on 24th September 2023. To only consider high fidelity entries compatible with the AlphaMissense data, the ClinVar data was filtered as follows: 1) Type == “single nucleotide variant”.

2) Assembly == ‘GRCh38’, 3) ReviewStatus must be one of the following “criteria provided, single submitter”, “criteria provided, multiple submitters, no conflicts” or “reviewed by expert panel”. To only extract pathogenic variants, the data was subsetted to variants with ClinicalSignificance being either of the following: ‘Likely pathogenic’, ‘Pathogenic/Likely pathogenic’, or ‘Pathogenic’. To integrate with the other datasets, we also created the “var_id” from the ClinVar data (see definition above). AlphaMissense predictions were then annotated as being in the ClinVar database by annotating if the constructed “var_id” were in both datasets, but this time also allowed for the reference and alternative nucleotide to be swapped as user-submitted data often mix these around (∼16.52% matched the reverse id).

#### Detecting isoform differences in variant interpretations

To analyze how different isoforms affect the interpretation of a variant, we first filtered for missense variants from genes where at least two isoforms were found (N_t-all_ >=2). We then, for each “var_id” summarized the information extracted above, allowing us to define 5 overlapping categories of isoform-dependent differences in variant interpretation: 1) “AlphaMissense Prediction “Benign” vs. “Pathogenic”” defined as a variant where the AlphaMissense prediction was “likely_benign” for one isoform but “likely_pathogenic” for another transcript. 2) “Different Missense Variants” were defined as cases where isoforms had different annotated amino acid variants. 3) “Missense vs Synonymous” defined by variants with N_t-synonyms_ > 0 (see definition above). 4) “Missense vs Non-coding” defined by variants with N_t-noncoding_ > 0 (see definition above). 5) “Variant Spliced Out” defined by variants with N_t-splice_ > 0 (see definition above). For the analysis, we then simply counted the occurrence of each category in the indicated subset of the data (subset defined above in respective settings), and fractions were extracted by dividing by the number of observed “var_id” in the corresponding category (after filtering indicated above).

### SpliceAI overlap

The SpliceAI[36] predictions of splice disrupting mutations were downloaded for every mutation at all positions in the human genome from https://basespace.illumina.com/s/otSPW8hnhaZR (the link is different from the original article because the original link expired. The authors provided this new one upon request). Specifically the “spliceai_scores.raw.snv.hg38.vcf.gz” file was downloaded, and we extracted positions where at least one of the 4 splice disruption scores (acceptor gain (AG), acceptor loss (AL), donor gain (DG), and donor loss (DL)) were >0.8 as this is the cutoff used to detect high-confidence splice-disruptions in the SpliceAI paper[36]. For variants passing the filter, we created the “var_id” (as definition above) and then counted the overlap of “var_id” between AlphaMissense variants predicted as “likely_pathogenic” and the filtered SpliceAI variants.

### Text augmentation

ChatGPT was used to aid in the creation of this manuscript. Specifically, we used ChatGPT v4o and v4.5 as an editor and asked the model to improve the text of specific sections we had trouble formulating to our satisfaction. All text was carefully read to ensure ChatGPT added no meaning-changing editions. ChatGPT and Perplexity.ai were also used, in addition to Pubmed and Google Scholar, to search for relevant references. All references were read and verified before being added to the manuscript.

## Results

### Overview of data analyzed

We first turned to RNA-seq-based QTL studies to investigate the importance of isoforms in the context of common genetic variants. We define cases in which a variant is associated with differential gene expression as eGenes. In contrast, cases in which a variant is associated with differential isoform usage are termed iuGenes (see Figure 1B for a schematic overview of these definitions). Importantly, by measuring the relative usage of isoforms, we can identify variants associated with changes in isoform composition independent of changes in the overall gene expression[37]. Next, we systematically extract significant eGenes and iuGenes from 83 RNA-seq-based QTL datasets (Figure 1C) covering 75 tissue- and cell types (Supplementary Table 2)[19] which we confirmed were of predominantly European ancestry (see methods, Supplementary Figure 1C.1).

### Systematic analysis of isoform usage QTLs

The number of significant iuGenes and eGenes was highly correlated (Pearson correlation: 0.92, *P*-value: 1.02e-34), and we found a median of 1,448 significant iuGenes and 1,666 significant eGenes (Figure 1D, Supplementary Data). Importantly, these results confirm and generalize recent tissue- or mechanism-specific findings using both short and long-read RNA-seq[38–43]. Although the number of significant eGenes and iuGenes was similar, we found that the overlap was minimal (median of 13.3%), and eGenes alone explain, on median, only 44.0% of the genes affected by common variants (Figure 1E). Interestingly, we find that among the genes affected by both gene-level and isoform usage-level changes, on median 81.3% have overlapping purity-filtered credible sets (Supplementary Figure 1E.1D), suggesting co-regulation of splicing and expression by the same variant. In summary words, isoform-level changes represent a major pathway through which common genetic variance influences the transcriptional output of the genome.

### Gene and isoform associations to GWAS phenotypes

We integrated the QTL data with thousands of GWAS datasets to analyze whether isoforms are also associated with human phenotypes. Specifically, we extracted all GWAS variants with genome-wide significance (*P*-value < 5e-8) from the EBI GWAS catalog[20] and used strict colocalization[24] to associate them with variants from significant eGenes and iuGenes. The results are specific associations, at both gene and isoform levels, between 9,691 GWAS phenotypes and 10,071 genes (Figure 1C, Supplementary Table 4, Supplementary Data). To analyze these associations, we initially focused on six disease-associated phenotypes that represent the primary causes of preventable mortality, diminished quality of life, and pressure on healthcare systems in developed countries (e.g., obesity, type-2 diabetes, cancer)[44]. We find that associations through overall gene expression alone only account for, on average, 59.0% of identified associations (Figure 1F). Notably, this finding generalizes across the 1,281 GWAS phenotypes that have at least 10 genes associated (median 53.9%, Figure 1G, Supplementary Figure 1G.1, Supplementary Table 4). This also means that 46.1% of the associations between GWAS phenotypes and the genes that enable mechanistic insights are affected at the isoform-level. Thus, isoform-level changes represent a fundamental mechanism by which genetic variation shapes human phenotypes in both health and disease.

### Genes utilize multiple isoforms

To address a potential criticism that isoform changes observed at the RNA level might not manifest at the protein level, we next performed a meta-analysis of proteomics data[3,32]. To test this hypothesis directly, we performed the first systematic meta-analysis of isoform abundance across numerous proteomics datasets. Specifically, we utilized a recent breakthrough that allows the quantification of individual isoforms from proteomics data[27] to analyze isoform usage from 56 proteomics studies (1,878 samples, Supplementary Table 5). We focused on protein-coding genes annotated with two or more isoforms in the GENCODE database[10]. For each sample and gene, we identified the most abundant isoform (often termed major isoform). We find that up to 32.8% of detected genes utilize multiple major isoforms across the datasets, thereby providing the first direct evidence of widespread differential isoform usage at the protein level (Figure 2A, Supplementary Figure 2A.1-2). Importantly, these findings cannot be explained by the effect of aggregation across many samples (Supplementary Figure 2A.3), the inherent uncertainty of assigning peptides to proteins[32] (Supplementary Figure 2A.4), or how a major isoform is defined (Supplementary Figure 2A.5). Furthermore, this finding is likely underestimated given that this data represents less than 0.5% of current human proteomics datasets and that the main driver of detected isoform diversity seems to be the number of datasets analyzed (Supplementary Figure 2A.6). Accordingly, we find that usage of multiple distinct major isoforms increases to 94.9% when we analyze 10,000+ human RNA-seq datasets[33] (Supplementary Figure 2A.7, Supplementary Table 6). Thus, through robust analysis of both proteomics and RNA-seq, we find that different isoforms are used in different tissues, cell types, and diseases and, hence, cannot be ignored when interpreting genetic variants.

### Variant interpretation in the context of isoforms

Given the evidence that almost half of genetic associations (Figure 1) and even protein abundance (Figure 2A) operate at the isoform level, we next asked if rare variant interpretations are similarly dependent on isoform context. A hypothetical example of such an isoform-specific effect could be a variant that affects only a specific tissue because other tissues use an isoform in which the variant exon is spliced out. Such examples of isoform-specific disease-causing variants exist in the literature[13–15], but it is unknown how common they are. To clarify this and thereby determine the size of the opportunity gap (Figure 1A), we next systematically investigated the importance of isoforms for rare variant analysis. Specifically, for all possible human missense variants (both predicted (n=∼76 million) and known (n=∼300,000)), we analyzed five different ways the interpretation of a variant can change depending on the underlying isoform (see schematic illustration in Figure 2C). Surprisingly, we find that, on average, 72.7% of analyzed human missense variants have a different functional interpretation in another isoform. Importantly, this finding holds true independently of whether we consider all possible missense variants, missense variants in known mutation databases (ClinVar[22], COSMIC[23]), missense variants predicted as pathogenic by AlphaMissense[21], or various combinations thereof (Figure 2D, Supplementary Figure 2D.1-2). Of particular note, we find that ∼3% of predicted pathogenic variants are predicted as benign in another protein-coding isoform – a number that is similar even when only considering variants from ClinVar or COSMIC (Figure 2D top-right). This means that a variant deemed benign without isoform context could, in fact, be pathogenic in a tissue that expresses an alternate isoform, and we find nearly 1 million such cases throughout the human genome (Supplementary Figure 2D.2). These findings are especially relevant given the widespread use of “reference transcript” catalogs such as MANE select[45] and APPRIS[46]. Although these resources are valuable, our results suggest that representing a gene with a single isoform may not fully capture the underlying biological complexity. Consistent with this, our analysis of RNA-seq data showed that transcripts not designated as “reference” by these resources were, on average, the most highly expressed (major) isoforms in more than 1,000 datasets (Supplementary Figure 2D.3), indicating that reference transcripts should be interpreted as practical representatives rather than universally dominant isoforms. Taken together, these results demonstrate that it is paramount to consider all isoforms when analyzing rare variants, as the interpretation of the variant almost always depends on the isoform expressed in the relevant tissue/cells.

However, the interpretation of coding variants is not the only way rare variants can affect isoforms. Variants affecting splice enhancer/silencer can change which isoforms are expressed[9]. Indeed, SpliceAI[36] finds more than 3 million such variants in the human genome that almost entirely differ from AlphaMissense Pathogenic predictions (Figure 2E). In addition, 41.9% of variants affecting RNA isoform-usage (iuQTL) are not located within the affected gene (neither exonic nor intronic) (Figure 2F, Supplementary Figure 2F.1). Jointly, this suggests that rare variants, both within and distal from affected genes, can change which isoforms are used and, hence both the interpretation of a variant or the biological function of the gene. In other words, isoform-level changes are essential when interpreting rare variants and might help explain a large proportion of current patients with undiagnosed diseases[47,48].

## Discussion

In summary, we have shown through systematic analysis that isoforms are crucial for understanding the effect of both common and rare genetic variants. From these results, it is clear that augmenting current genetic analysis of gene-level data with isoform-level analysis leads to improved functional understanding[49], enables the discovery of many otherwise overlooked affected genes (Figure 1E, 1G), and offers new opportunities and challenges for drug development[9,50,51]. The high correlation of eGenes and iuGenes (Figure 1D) suggests that in other QTL studies where eGenes are identified, we would also expect to find many iuGenes, suggesting widespread reanalysis of genetics studies could lead to additional biological insights. This argument is further enhanced by recent findings suggesting that isoform-level analysis even improves the detection of gene-level effects[52].

While we find that isoform-level analysis of genetics data is paramount, our results underestimate the true importance of isoform-level changes. This underestimation is partly because statistical analysis of isoforms is almost always underpowered compared to gene-level analysis[53] and partly because the analysis pipeline used here does not consider isoform-expression QTLs, novel-splice sites, or novel isoforms[19]. The latter is emphasized by a recent long-read QTL study identifying thousands of novel isoforms regulated by common genetic variants[43]. These long-read-based analyses, however, also highlight a limitation of our study: our analysis relies on isoform quantification of short-read RNA-seq, which are sensitive to annotation quality and model assumptions, especially for lowly expressed or highly similar isoforms.

The importance of our results is further highlighted by our analysis of proteomics data, which provides the first genome-wide direct evidence of widespread differential protein isoform abundance, thereby moving isoform-level evidence beyond splice-site-based proteomics QTL studies[41,54–56]. The omnipresence of isoform importance in genetics also highlights the need to understand the biological function of isoforms. However, since all current bioinformatic tools that enable functional analysis of isoforms (e.g., IsoformSwitchAnalyzeR[57] and tappAS[58]) rely on sequence-based annotation, it is clear there is an acute need for novel bioinformatic tools and databases that enable biological interpretation of isoform level changes.

Taken together, the work presented here argues that moving from gene-level to isoform-resolved genetics could improve the interpretation of clinical variants, because variants that appear benign or ambiguous at the gene level may, in fact, disrupt the disease-relevant transcript in a tissue- or context-specific manner or vice versa. That is directly relevant to rare-disease diagnostics and VUS reclassification, where transcript-aware interpretation and RNA evidence are increasingly recognized as necessary for assessing splice-disrupting and isoform-specific pathogenic variants[59]. This is further supported by our finding that the vast majority (79.8%) of co-occurring eQTL and iuQTL signals at the same gene are driven by a single shared causal variant, underscoring the mechanistic importance of isoform-resolved analysis.

The implications for drug development are similarly important. Human genetic analyses of both common and rare variation are increasingly used to identify and prioritize drug targets[60], and targets with genetic support are more likely to succeed during clinical development[61]. In this context, our results suggest that isoform-level analysis is both a challenge and an opportunity: a challenge because current analytical infrastructure still largely operates at the gene level, but an opportunity because, across diseases, isoform-resolved analyses identified on average 25% additional genes beyond gene-level approaches (Figure 1F,G), thereby expanding the pool of candidate therapeutic targets and highlighting the value of retrospective isoform-resolved reanalysis of existing genetic datasets. This is especially interesting since it not only gives rise to a new class of drugs that specifically target isoforms, but also because such drugs might be safer for patients to use[62].

While all results here are based on aggregation across all cells/tissue/genes/traits to present a global overview, we recognize that many researchers are interested in specific genes/traits in select tissues. Therefore, we have developed an <u>R Shiny app</u> that lets the scientific community explore individual genes/traits in a tissue/cell-type-specific manner, inspect associated isoforms and GWAS traits, view genomic context with transcript structures and external Ensembl/UCSC/GeneCards links, download result tables, and generate shareable bookmarked URLs for selected genes or traits. Furthermore, all intermediate results are available online via <u>Zenodo</u>, enabling the community to perform systematic analyses of isoforms, e.g., in a tissue-specific manner.

### Conclusion

Our analysis of common variants suggests that almost half of human genes (Figure 1E.1) and phenotypes (Figure 1F-G) affected by genetic variation are affected at the isoform level. In other words, this adds a second dimension to the current gene-centric analysis approach, thereby providing a more holistic picture of human genetics. This aligns with our recent large-scale reanalysis of RNA-seq studies, which shows that, on average, 48.1% of the biologically meaningful transcriptomic signal involves isoform-level regulation [11]. Taken together, this suggests that, regardless of the organizational level analyzed, isoform-level changes are as relevant as gene-level changes (DNA: Figure 1D; RNA: Dam *et al.*^11^; Proteins: Figure 2A; Human Phenotypes: Figure 1F-G). Given that only a minority of contemporary studies consider isoforms (Figure 1A and reference[11]), we believe isoform-level analysis holds a significant promise for improving our understanding of the human condition in both health and disease.

## Declarations Acknowledgments

Thanks to the LDlinkR team at NIH for granting us VIP access to using the LDlinkR server. We want to thank Laura Maarit Pikkupeura, Frederik Otzen Bagger and Gloria Sheynkman for their insightful feedback on the manuscript.

## Authors’ contributions

KVS conceived the study. CH conducted the literature survey and curated the GWAS data. JMM, DSK and KVS analyzed and interpreted all data except for the proteomics-based analysis, which was conducted by KVS and ST. DSK performed eQTL dataset ancestry validation, eQTL/iuQTL overlap and credible-set distance analyses and developed the Shiny app. P.W.S. assisted with Shiny application debugging, deployment, and server-side analysis support. JMM, DSK and KVS wrote the manuscript. All authors read and approved the final manuscript.

## Consent for publication

Not applicable.

## Ethics approval and consent to participate

Not applicable. This study is a re-analysis of publicly available data and does not involve human subjects’ research.

## Availability of data and materials

All QTL datasets were obtained from the EBI QTL Catalogue (https://www.ebi.ac.uk/eqtl/). GWAS associations were obtained from the NHGRI-EBI GWAS Catalog (https://www.ebi.ac.uk/gwas/). Proteomics datasets were obtained from PRIDE (https://www.ebi.ac.uk/pride/; accessions in Supplementary Table 5), and RNA-seq datasets used for dominant-isoform analyses were obtained from ARCHS4 (https://maayanlab.cloud/archs4/; GEO accessions in Supplementary Table 6). AlphaMissense predictions were obtained from Zenodo (https://zenodo.org/records/8208688), ClinVar annotations from NCBI ClinVar (https://ftp.ncbi.nlm.nih.gov/pub/clinvar/tab_delimited/), COSMIC data from COSMIC (https://cancer.sanger.ac.uk/cosmic/), and SpliceAI predictions from Illumina BaseSpace (https://basespace.illumina.com/s/otSPW8hnhaZR). APPRIS annotations were obtained from https://appris.bioinfo.cnio.es/#/downloads, and MANE Select annotations from https://ftp.ncbi.nlm.nih.gov/refseq/MANE/MANE_human/release_1.5/. Processed results and Shiny app input data are available through Zenodo record 10.5281/zenodo.18862578 (https://zenodo.org/records/18862578), and the interactive Shiny app is available at https://services.healthtech.dtu.dk/shiny/qtl_gwas/.

## Supporting information

Supplementary_Table1_literature_review

Supplementary_Table3_ancestry

Supplementary_Table4_GWAS_trait_annotation

Supplementary_Table5_proteomics

Supplementary_Table6_RNA_seq

Supplementary_Table7_gene_overlap

Supplementary_Table8_variant_overlap

## Data Availability

All data produced in the present work are contained in the manuscript and its supplementary tables and Zenodo.

https://zenodo.org/records/15106238

## References

1. Cheng A, Harikrishna JA, Redwood CS, Lit LC, Nath SK, Chua KH. Genetics Matters: Voyaging from the Past into the Future of Humanity and Sustainability. Int J Mol Sci. 2022;23:3976. 10.3390/ijms23073976

2. Rockman MV, Kruglyak L. Genetics of global gene expression. Nat Rev Genet. 2006;7:862–72. 10.1038/nrg1964

3. Wright CJ, Smith CWJ, Jiggins CD. Alternative splicing as a source of phenotypic diversity. Nat Rev Genet. 2022;23:697–710. 10.1038/s41576-022-00514-4

4. Manolio TA, Collins FS, Cox NJ, Goldstein DB, Hindorff LA, Hunter DJ, et al. Finding the missing heritability of complex diseases. Nature. 2009;461:747–53. 10.1038/nature08494

5. Uffelmann E, Huang QQ, Munung NS, Vries J de, Okada Y, Martin AR, et al. Genome-wide association studies. Nat Rev Methods Prim. 2021;1:59. 10.1038/s43586-021-00056-9

6. Momozawa Y, Mizukami K. Unique roles of rare variants in the genetics of complex diseases in humans. J Hum Genet. 2021;66:11–23. 10.1038/s10038-020-00845-2

7. Schork NJ, Murray SS, Frazer KA, Topol EJ. Common vs. rare allele hypotheses for complex diseases. Curr Opin Genet Dev. 2009;19:212–9. 10.1016/j.gde.2009.04.010

8. Chen W, Coombes BJ, Larson NB. Recent advances and challenges of rare variant association analysis in the biobank sequencing era. Front Genet. 2022;13:1014947. 10.3389/fgene.2022.1014947

9. Marasco LE, Kornblihtt AR. The physiology of alternative splicing. Nat Rev Mol Cell Bio. 2023;24:242–54. 10.1038/s41580-022-00545-z

10. Frankish A, Carbonell-Sala S, Diekhans M, Jungreis I, Loveland JE, Mudge JM, et al. GENCODE: reference annotation for the human and mouse genomes in 2023. Nucleic Acids Res. 2022;51:D942–9. 10.1093/nar/gkac1071

11. Dam SH, Olsen LR, Vitting-Seerup K. Expression and splicing mediate distinct biological signals. BMC Biol. 2023;21:220. 10.1186/s12915-023-01724-w

12. Schwerk C, Schulze-Osthoff K. Regulation of Apoptosis by Alternative Pre-mRNA Splicing. Mol Cell. 2005;19:1–13. 10.1016/j.molcel.2005.05.026

13. Cummings BB, Karczewski KJ, Kosmicki JA, Seaby EG, Watts NA, Singer-Berk M, et al. Transcript expression-aware annotation improves rare variant interpretation. Nature. 2020;581:452–8. 10.1038/s41586-020-2329-2

14. Guven A, Tolun A. TBC1D24 truncating mutation resulting in severe neurodegeneration. J Méd Genet. 2013;50:199. 10.1136/jmedgenet-2012-101313

15. Nousbeck J, Burger B, Fuchs-Telem D, Pavlovsky M, Fenig S, Sarig O, et al. A Mutation in a Skin-Specific Isoform of SMARCAD1 Causes Autosomal-Dominant Adermatoglyphia. Am J Hum Genet. 2011;89:302–7. 10.1016/j.ajhg.2011.07.004

16. Kwan T, Benovoy D, Dias C, Gurd S, Provencher C, Beaulieu P, et al. Genome-wide analysis of transcript isoform variation in humans. Nat Genet. 2008;40:225–31. 10.1038/ng.2007.57

17. Consortium TGte. The GTEx Consortium atlas of genetic regulatory effects across human tissues. Science. 2020;369:1318–30. 10.1126/science.aaz1776

18. Li YI, Geijn B van de, Raj A, Knowles DA, Petti AA, Golan D, et al. RNA splicing is a primary link between genetic variation and disease. Science. 2016;352:600–4. 10.1126/science.aad9417

19. Kerimov N, Hayhurst JD, Peikova K, Manning JR, Walter P, Kolberg L, et al. A compendium of uniformly processed human gene expression and splicing quantitative trait loci. Nat Genet. 2021;53:1290–9. 10.1038/s41588-021-00924-w

20. Sollis E, Mosaku A, Abid A, Buniello A, Cerezo M, Gil L, et al. The NHGRI-EBI GWAS Catalog: knowledgebase and deposition resource. Nucleic Acids Res. 2022;51:D977–85. 10.1093/nar/gkac1010

21. Cheng J, Novati G, Pan J, Bycroft C, Žemgulytė A, Applebaum T, et al. Accurate proteome-wide missense variant effect prediction with AlphaMissense. Science. 2023;381:eadg7492. 10.1126/science.adg7492

22. Landrum MJ, Lee JM, Riley GR, Jang W, Rubinstein WS, Church DM, et al. ClinVar: public archive of relationships among sequence variation and human phenotype. Nucleic Acids Res. 2014;42:D980–5. 10.1093/nar/gkt1113

23. Sondka Z, Dhir NB, Carvalho-Silva D, Jupe S, Madhumita, McLaren K, et al. COSMIC: a curated database of somatic variants and clinical data for cancer. Nucleic Acids Res. 2023;52:D1210–7. 10.1093/nar/gkad986

24. Myers TA, Chanock SJ, Machiela MJ. LDlinkR: An R Package for Rapidly Calculating Linkage Disequilibrium Statistics in Diverse Populations. Front Genet. 2020;11:157. 10.3389/fgene.2020.00157

25. Consortium Gte, Gamazon ER, Segrè AV, Bunt M van de, Wen X, Xi HS, et al. Using an atlas of gene regulation across 44 human tissues to inform complex disease- and trait-associated variation. Nat Genet. 2018;50:956–67. 10.1038/s41588-018-0154-4

26. Colocalization of GWAS and eQTL signals at loci with multiple signals identifies additional candidate genes for body fat distribution - PMC [Internet]. [cited 2026 Apr 13]. https://pmc.ncbi.nlm.nih.gov/articles/PMC7202621/?utm_source=chatgpt.com. Accessed 13 Apr 2026

27. Bollon J, Shortreed MR, Jordan BT, Miller R, Jeffery E, Cavalli A, et al. IsoBayes: a Bayesian approach for single-isoform proteomics inference. bioRxiv. 2024;2024.06.10.598223. 10.1101/2024.06.10.598223

28. Cox J, Hein MY, Luber CA, Paron I, Nagaraj N, Mann M. Accurate Proteome-wide Label-free Quantification by Delayed Normalization and Maximal Peptide Ratio Extraction, Termed MaxLFQ*. Mol Cell Proteomics. 2014;13:2513–26. 10.1074/mcp.m113.031591

29. Perez-Riverol Y, Bandla C, Kundu DJ, Kamatchinathan S, Bai J, Hewapathirana S, et al. The PRIDE database at 20 years: 2025 update. Nucleic Acids Res. 2024;53:D543–53. 10.1093/nar/gkae1011

30. Deutsch EW, Overall CM, Eyk JEV, Baker MS, Paik Y-K, Weintraub ST, et al. Human Proteome Project Mass Spectrometry Data Interpretation Guidelines 2.1. J Proteome Res. 2016;15:3961–70. 10.1021/acs.jproteome.6b00392

31. Webel H, Perez-Riverol Y, Nielsen AB, Rasmussen S. Mass spectrometry-based proteomics data from thousands of HeLa control samples. Sci Data. 2024;11:112. 10.1038/s41597-024-02922-z

32. Tress ML, Abascal F, Valencia A. Alternative Splicing May Not Be the Key to Proteome Complexity. Trends Biochem Sci. 2017;42:98–110. 10.1016/j.tibs.2016.08.008

33. Lachmann A, Torre D, Keenan AB, Jagodnik KM, Lee HJ, Wang L, et al. Massive mining of publicly available RNA-seq data from human and mouse. Nat Commun. 2018;9:1366. 10.1038/s41467-018-03751-6

34. Bray NL, Pimentel H, Melsted P, Pachter L. Near-optimal probabilistic RNA-seq quantification. Nat Biotechnol. 2016;34:525–7. 10.1038/nbt.3519

35. Lawrence M, Huber W, Pagès H, Aboyoun P, Carlson M, Gentleman R, et al. Software for Computing and Annotating Genomic Ranges. PLoS Comput Biol. 2013;9:e1003118. 10.1371/journal.pcbi.1003118

36. Jaganathan K, Panagiotopoulou SK, McRae JF, Darbandi SF, Knowles D, Li YI, et al. Predicting Splicing from Primary Sequence with Deep Learning. Cell. 2019;176:535–548.e24. 10.1016/j.cell.2018.12.015

37. Vitting-Seerup K, Sandelin A. The Landscape of Isoform Switches in Human Cancers. Mol Cancer Res. 2017;15:1206–20. 10.1158/1541-7786.mcr-16-0459

38. Brotman SM, Raulerson CK, Vadlamudi S, Currin KW, Shen Q, Parsons VA, et al. Subcutaneous adipose tissue splice quantitative trait loci reveal differences in isoform usage associated with cardiometabolic traits. Am J Hum Genet. 2022;109:66–80. 10.1016/j.ajhg.2021.11.019

39. Qi T, Wu Y, Fang H, Zhang F, Liu S, Zeng J, et al. Genetic control of RNA splicing and its distinct role in complex trait variation. Nat Genet. 2022;1–9. 10.1038/s41588-022-01154-4

40. Bhattacharya A, Vo DD, Jops C, Kim M, Wen C, Hervoso JL, et al. Isoform-level transcriptome-wide association uncovers genetic risk mechanisms for neuropsychiatric disorders in the human brain. Nat Genet. 2023;55:2117–28. 10.1038/s41588-023-01560-2

41. Abood A, Mesner LD, Jeffery ED, Murali M, Lehe MD, Saquing J, et al. Long-read proteogenomics to connect disease-associated sQTLs to the protein isoform effectors of disease. Am J Hum Genet. 2024;111:1914–31. 10.1016/j.ajhg.2024.07.003

42. Raj T, Li YI, Wong G, Humphrey J, Wang M, Ramdhani S, et al. Integrative transcriptome analyses of the aging brain implicate altered splicing in Alzheimer’s disease susceptibility. Nat Genet. 2018;50:1584–92. 10.1038/s41588-018-0238-1

43. Nagura Y, Shimada M, Kuribayashi R, Kiyose H, Igarashi A, Kaname T, et al. Long-read sequencing reveals novel isoform-specific eQTLs and regulatory mechanisms of isoform expression. medRxiv. 2024;2024.02.21.24302494. 10.1101/2024.02.21.24302494

44. Collaborators G 2021 C of D, Naghavi M, Ong KL, Aali A, Ababneh HS, Abate YH, et al. Global burden of 288 causes of death and life expectancy decomposition in 204 countries and territories and 811 subnational locations, 1990–2021: a systematic analysis for the Global Burden of Disease Study 2021. Lancet. 2024;403:2100–32. 10.1016/s0140-6736(24)00367-2

45. Morales J, Pujar S, Loveland JE, Astashyn A, Bennett R, Berry A, et al. A joint NCBI and EMBL-EBI transcript set for clinical genomics and research. Nature. 2022;604:310–5. 10.1038/s41586-022-04558-8

46. Pozo F, Rodriguez JM, Gómez LM, Vázquez J, Tress ML. APPRIS principal isoforms and MANE Select transcripts define reference splice variants. Bioinformatics. 2022;38:ii89–94. 10.1093/bioinformatics/btac473

47. Lord J, Baralle D. Splicing in the Diagnosis of Rare Disease: Advances and Challenges. Front Genet. 2021;12:689892. 10.3389/fgene.2021.689892

48. Blakes AJM, Wai HA, Davies I, Moledina HE, Ruiz A, Thomas T, et al. A systematic analysis of splicing variants identifies new diagnoses in the 100,000 Genomes Project. Genome Med. 2022;14:79. 10.1186/s13073-022-01087-x

49. Grarup N, Moltke I, Andersen MK, Dalby M, Vitting-Seerup K, Kern T, et al. Loss-of-function variants in ADCY3 increase risk of obesity and type 2 diabetes. Nat Genet. 2018;50:172–4. 10.1038/s41588-017-0022-7

50. Ji Y, Mishra RK, Davuluri RV. In silico analysis of alternative splicing on drug-target gene interactions. Sci Rep-uk. 2020;10:134. 10.1038/s41598-019-56894-x

51. Monteys AM, Hundley AA, Ranum PT, Tecedor L, Muehlmatt A, Lim E, et al. Regulated control of gene therapies by drug-induced splicing. Nature. 2021;596:291–5. 10.1038/s41586-021-03770-2

52. LaPierre N, Pimentel H. Accounting for isoform expression increases power to identify genetic regulation of gene expression. PLOS Comput Biol. 2024;20:e1011857. 10.1371/journal.pcbi.1011857

53. Love MI, Soneson C, Patro R. Swimming downstream: statistical analysis of differential transcript usage following Salmon quantification. F1000research. 2018;7:952. 10.12688/f1000research.15398.1

54. Mirauta BA, Seaton DD, Bensaddek D, Brenes A, Bonder MJ, Kilpinen H, et al. Population-scale proteome variation in human induced pluripotent stem cells. eLife. 2020;9:e57390. 10.7554/elife.57390

55. Suhre K, Chen Q, Halama A, Mendez K, Dahlin A, Stephan N, et al. A genome-wide association study of mass spectrometry proteomics using a nanoparticle enrichment platform. Nat Genet. 2025;57:2987–96. 10.1038/s41588-025-02413-w

56. Han Y, Wood SD, Wright JM, Dostal V, Lau E, Lam MPY. Computation-assisted targeted proteomics of alternative splicing protein isoforms in the human heart. J Mol Cell Cardiol. 2021;154:92–6. 10.1016/j.yjmcc.2021.01.007

57. Vitting-Seerup K, Sandelin A. IsoformSwitchAnalyzeR: analysis of changes in genome-wide patterns of alternative splicing and its functional consequences. Bioinformatics. 2019;35:4469–71. 10.1093/bioinformatics/btz247

58. Fuente L de la, Arzalluz-Luque Á, Tardáguila M, Risco H del, Martí C, Tarazona S, et al. tappAS: a comprehensive computational framework for the analysis of the functional impact of differential splicing. Genome Biol. 2020;21:119. 10.1186/s13059-020-02028-w

59. Truty R, Ouyang K, Rojahn S, Garcia S, Colavin A, Hamlington B, et al. Spectrum of splicing variants in disease genes and the ability of RNA analysis to reduce uncertainty in clinical interpretation. Am J Hum Genet. 2021;108:696–708. 10.1016/j.ajhg.2021.03.006

60. Plenge RM, Scolnick EM, Altshuler D. Validating therapeutic targets through human genetics. Nat Rev Drug Discov. 2013;12:581–94. 10.1038/nrd4051

61. Nelson MR, Tipney H, Painter JL, Shen J, Nicoletti P, Shen Y, et al. The support of human genetic evidence for approved drug indications. Nat Genet. 2015;47:856–60. 10.1038/ng.3314

62. Kjer-Hansen P, Phan TG, Weatheritt RJ. Protein isoform-centric therapeutics: expanding targets and increasing specificity. Nat Rev Drug Discov. 2024;23:759–79. 10.1038/s41573-024-01025-z

