## Supplementary figures for "Beyond the Gene in Genetics: How Isoform-Resolved Analysis Empowers the Study of Both Common and Rare Genetic Variation"

**Please note**: The figure numbering used here is not consecutive numbering but instead directly associates the supplementary figure with the main figure panel by simply using the main figure reference (e.g., "Figure 1G") followed by a ".X" where X is a number (since a main panel can have multiple supplementary figures, e.g., "Figure 1G.1"). These will naturally be changed to consecutive numbering after all revisions are done. Still, until that point, this approach minimizes the editing needed when main or supplementary figures are added/removed.

### Figure 1

#### *Figure 1C.1*

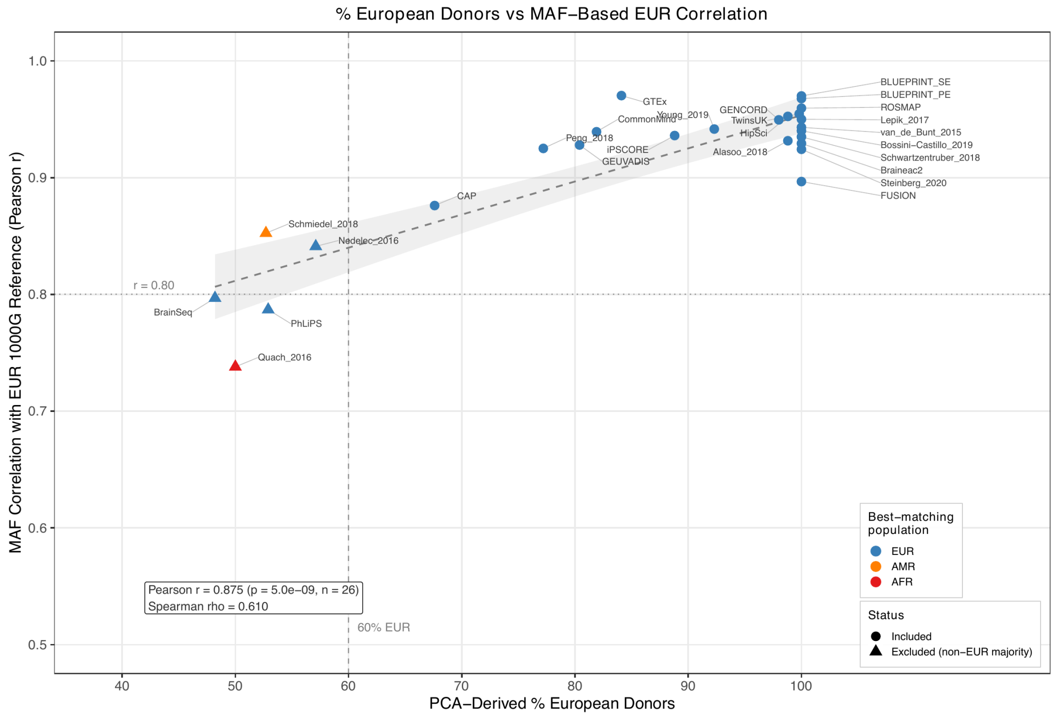

Pearson correlation between PCA-derived percentage of European donors (x-axis) and the correlation between study-level QTL minor allele frequencies and the 1000 Genomes EUR reference minor allele frequencies (y-axis) across 26 parent studies. Each point represents one parent study, with color indicating the best-matching 1000 Genomes superpopulation based on MAF correlation and shape indicating whether the study was retained or excluded. The vertical dashed line indicates the 60% EUR donor cutoff used for exclusion, and the horizontal dotted line marks MAF correlation r = 0.80 as a visual reference. Pearson r = 0.875, *P*-value = 5e-9; Spearman rho = 0.610. See Supplementary Table 3.

#### *Figure 1E*

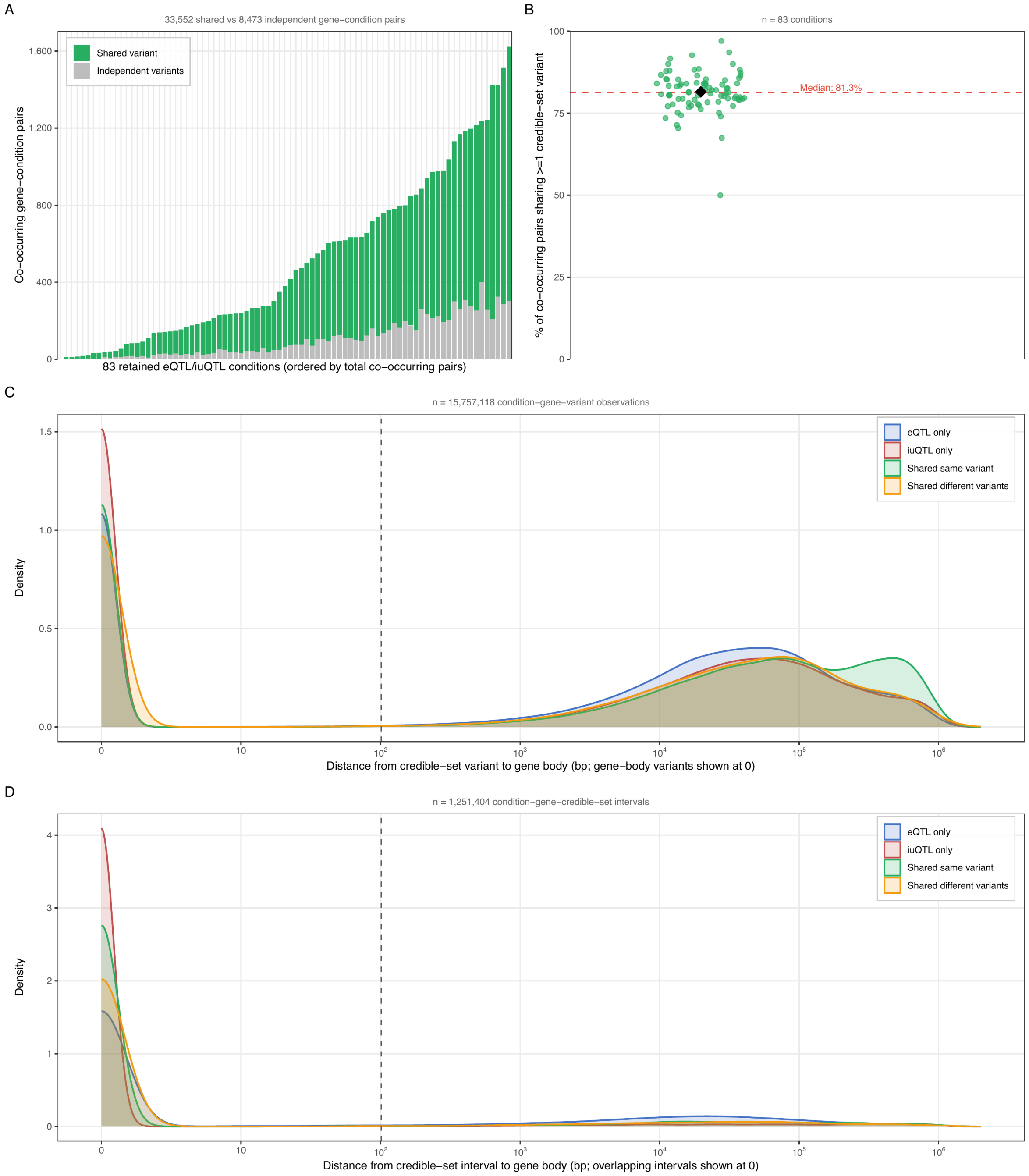

Gene- and variant-level overlap between eQTL and iuQTL signals across 83 retained paired eQTL/iuQTL conditions. A) Condition-level counts of co-occurring gene-condition pairs sharing at least one credible-set variant versus independently regulated pairs. B) Percentage of co-occurring gene-condition pairs sharing at least one credible-set variant across the 83 retained conditions (median 81.3%). C) Density of distances from credible-set variants to their associated QTL gene bodies by gene-condition category (n = 15,757,118 condition-gene-variant observations). D) Density of distances from credible-set interval spans to their associated QTL gene bodies for the same categories (n = 1,251,404 condition-gene-credible-set intervals). Distances are zero for gene-body or overlapping observations and are plotted as distance + 1 on a log10 scale. Overall, 33,552 of 42,025 co-occurring gene-condition pairs (79.8%) share at least one credible-set variant, while 8,473 pairs (20.2%) show independent eQTL and iuQTL signals. See Supplementary Tables 7 and 8.

*Figure 1G.1*

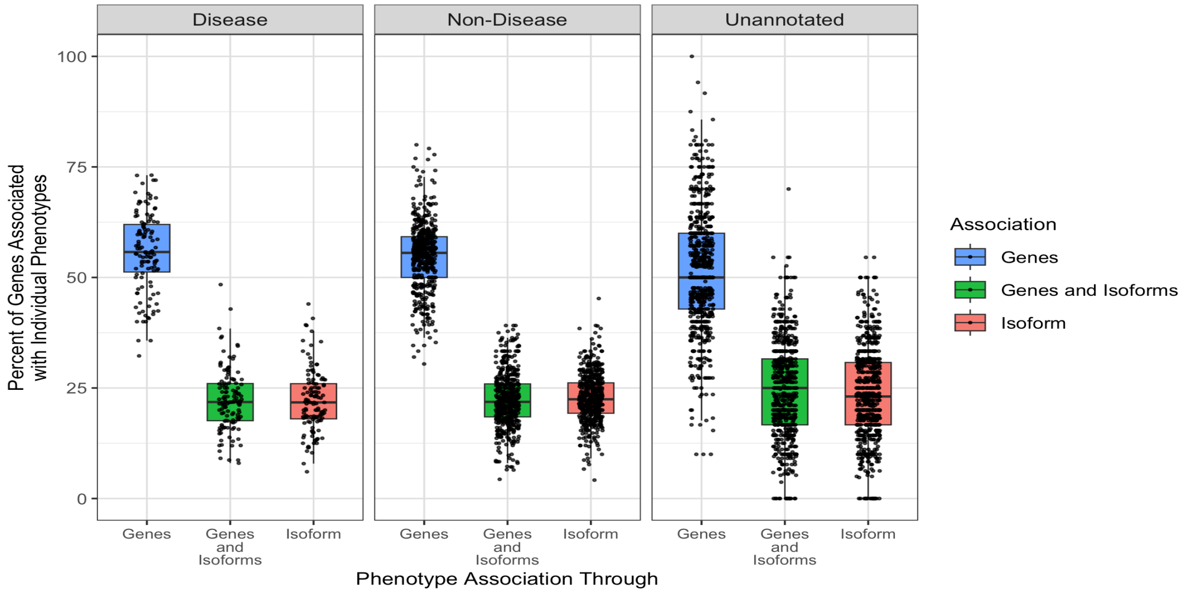


For 1,281 GWAS phenotypes (dots), the percent of colocalized genes (y-axis) is shown with color and x-axis indicating the QTL type giving rise to the association. Each phenotype has at least 10 colocalized genes, and subplots indicate the manual annotation of the GWAS phenotype. Gene expression alone explains a median of 53.9% of gene associations, indicating that isoform-level associations contribute broadly across both disease and non-disease GWAS phenotypes. See Supplementary Table 4.

### Figure 2

*Figure 2A.1*

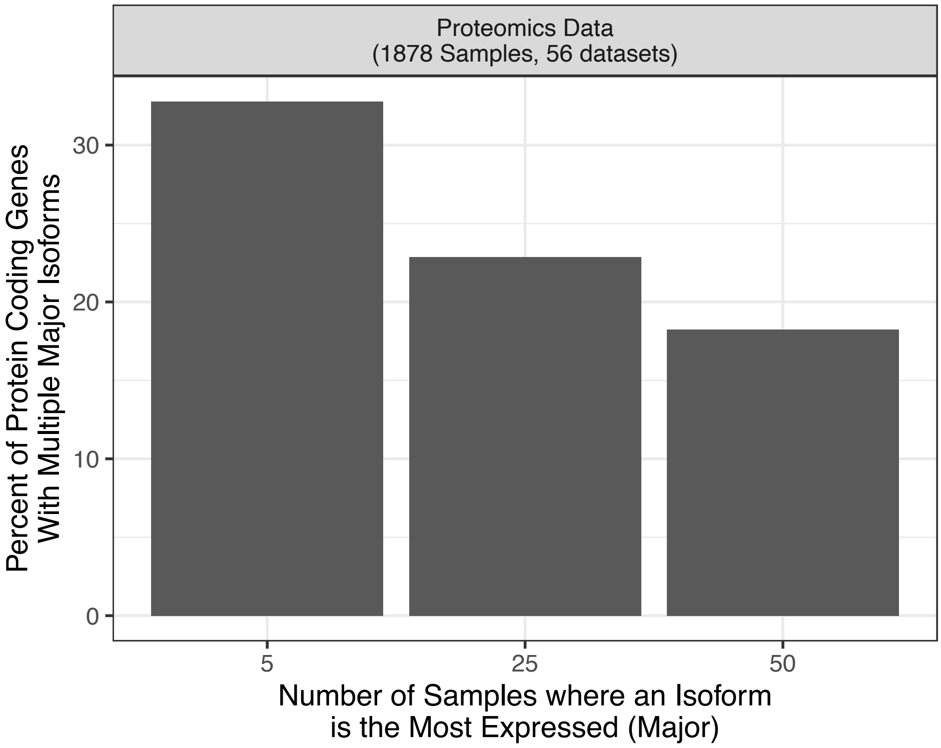


The percent of multi-isoform protein-coding genes (y-axis) with at least two major isoforms plotted against the minimum number of samples where an isoform had to be detected as the most expressed (Major). This shows that for 1000-2000 genes, we can confidently detect multiple distinct protein isoforms.

*Figure 2A.2*

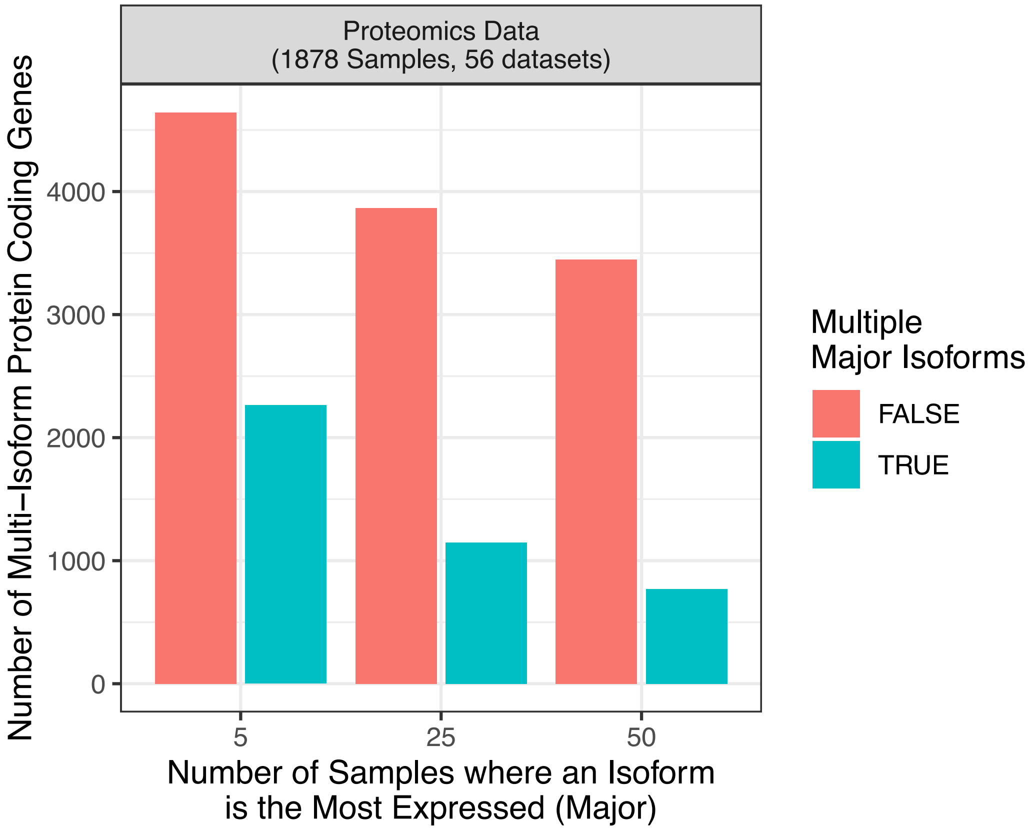

The number of detected multi-isoform protein-coding genes (y-axis) with at least two major isoforms (color) plotted against the minimum number of samples where an isoform had to be detected as the most expressed (Major). This shows that for 1000-2000 genes, we can confidently detect multiple distinct protein isoforms.

*Figure 2A.3*

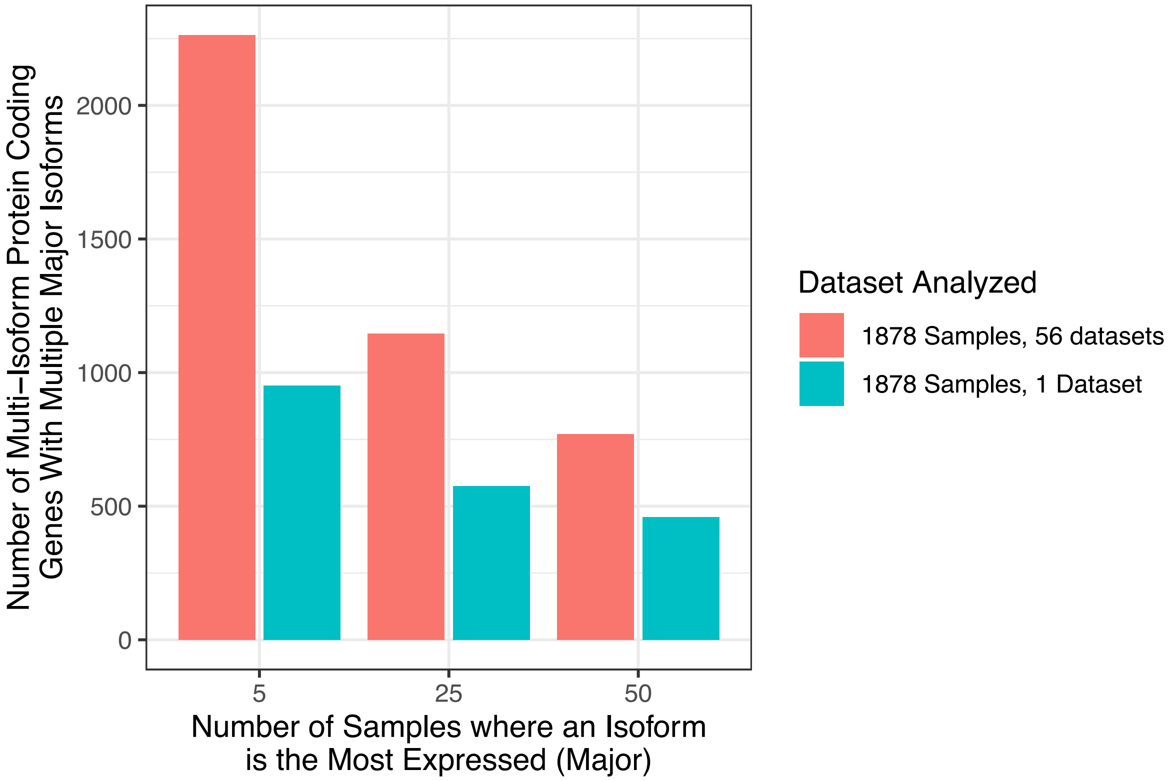


The number of detected multi-isoform protein-coding genes with at least two major isoforms (y-axis) plotted against the minimum number of samples where an isoform had to be detected as the most expressed (Major). Color shows two datasets analyzed: The collection of 56 proteomics datasets (1878 samples, red) used in the main figure and a separate single proteomics dataset with an identical sample size (blue). This shows that the number of samples analyzed is not the primary driver of detected multi-isoform genes.

*Figure 2A.4*

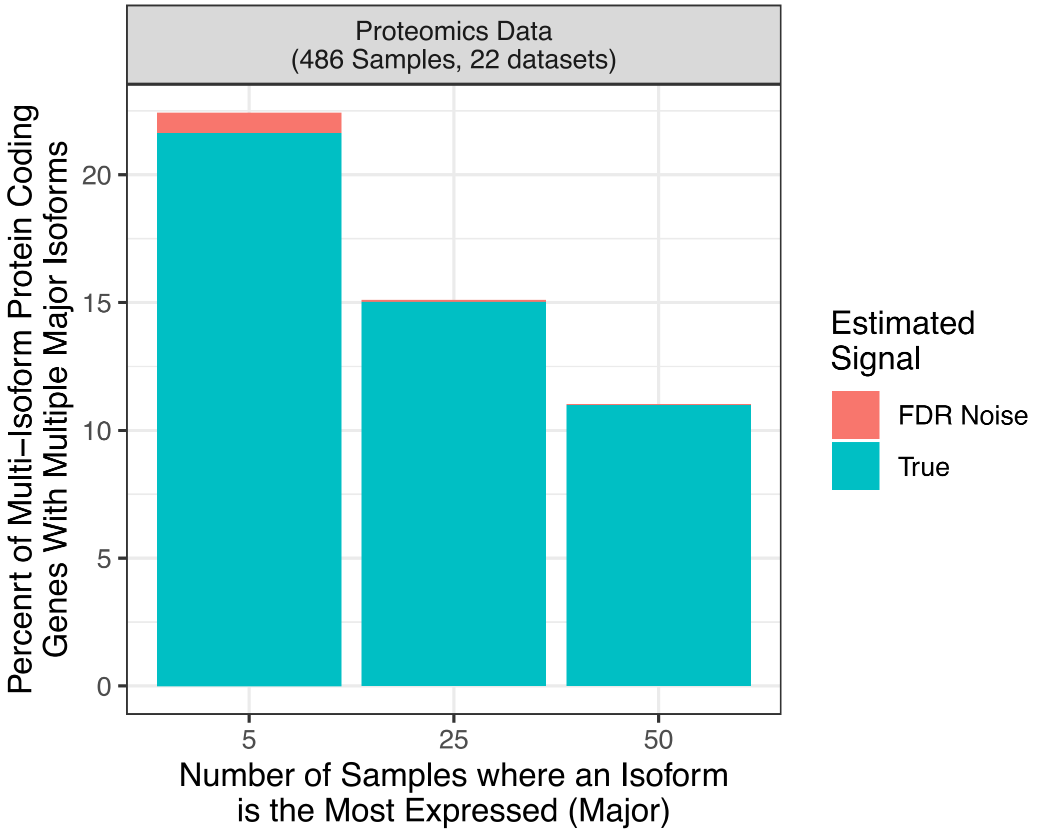


One of the major concerns for isoform analysis of proteomics data is that each peptide, due to how MS/MS signals are analyzed, has a slight chance (1% FDR) of being wrongly assigned. This naturally has the potential to detect isoforms wrongly, and hence, we investigated the importance of this potential error using a subset of the entire dataset (486 Samples, 22 datasets, see methods). Briefly, for each sample, we randomly re-assigned 1% of peptides to another annotated isoform (and thereby most likely also gene). We then re-quantified the data using Isobayes and repeated the analysis, counting genes with multiple major isoforms. The re-assignment and re-quantification were repeated 20 times, and the median percent detected multi-isoform was calculated. The FDR noise was then defined as the median percent (w added peptide scrambling) subtracted from the observed fraction, and that fraction is indicated as FDR noise by color. The plot shows the percent protein-coding genes with at least two major isoforms (y-axis) plotted against the minimum number of samples where an isoform had to be detected as the most expressed (Major). This analysis shows that the 1% FDR noise in the peptide data only explains a tiny proportion of the observed multi-isoform protein-coding genes, adding to the robustness of the result presented in the main figure.

*Figure 2A.5*

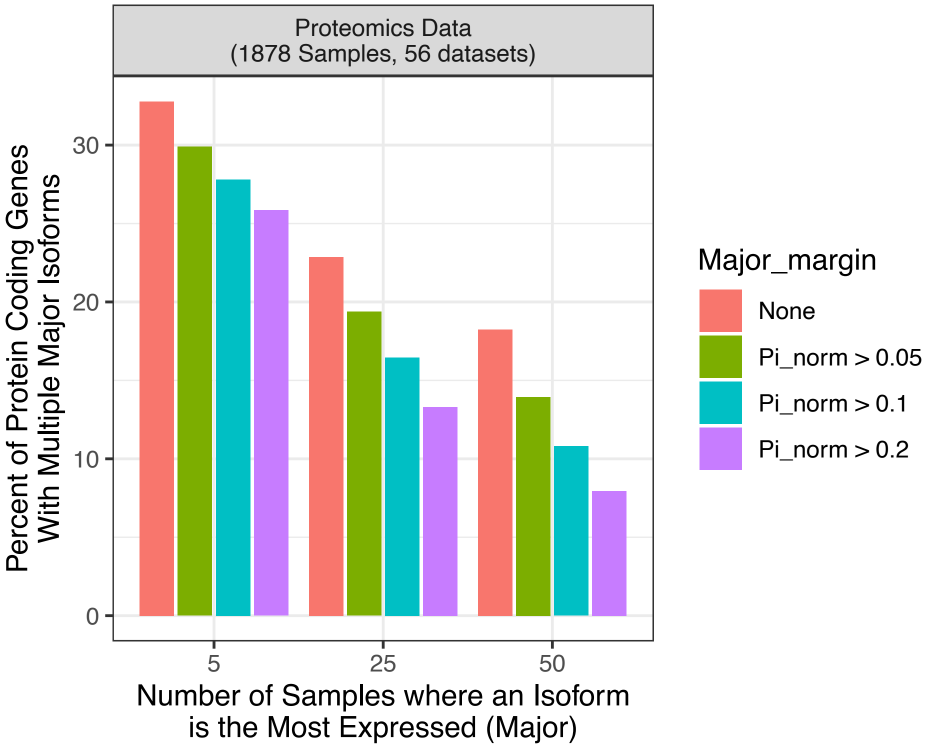


The percent of multi-isoform protein-coding genes (y-axis) with at least two major isoforms plotted against the minimum number of samples where an isoform had to be detected as the most expressed (Major). Color shows the effect of adding a requirement that the major isoform contributes 5%, 10% or 20% more to the total protein abundance than the second most abundant isoform. This shows that equally abundant isoforms do not contribute a lot to rates of detecting genes with multiple major isoforms. Please note that the stringent requirements also mean that a lot of genes which actually have multiple major isoforms will now be missed.

*Figure 2A.6*

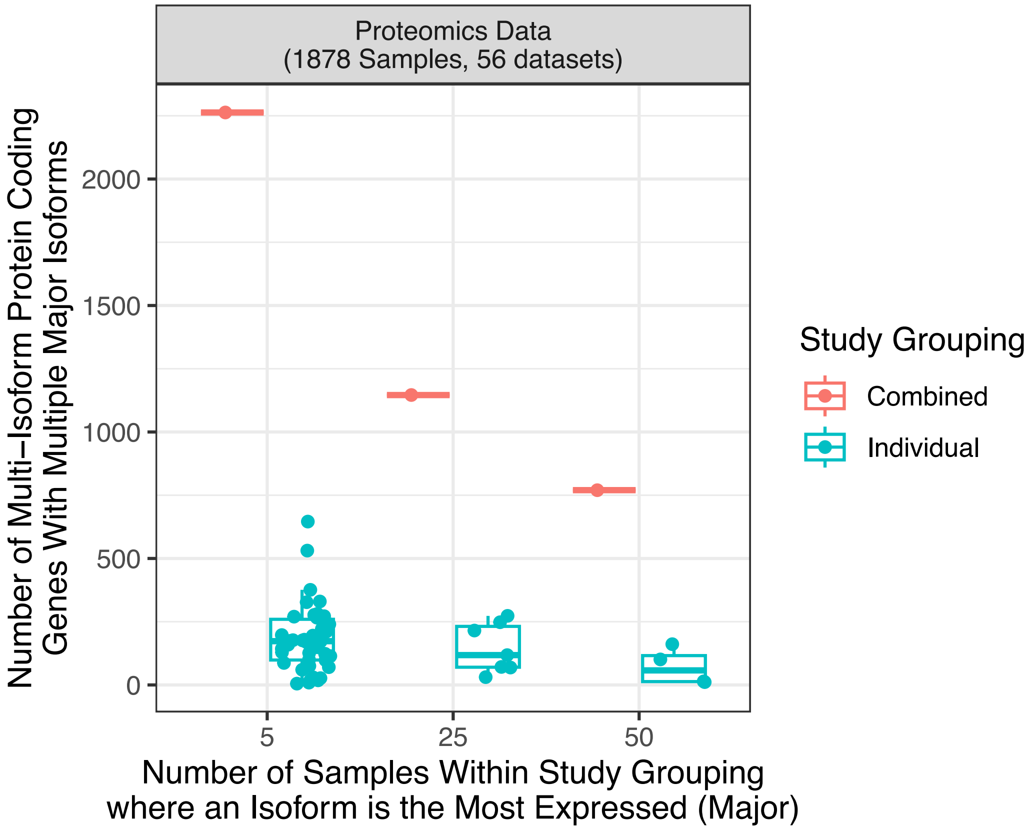


The number of detected multi-isoform protein-coding genes (y-axis) with at least two major isoforms is plotted against the minimum number of samples where an isoform had to be detected as the most expressed (Major). Each dot is either an individual dataset (blue) or the combined dataset (red). This indicates that the main power of this analysis comes from aggregating across individual proteomics datasets.

*Figure 2A.7*

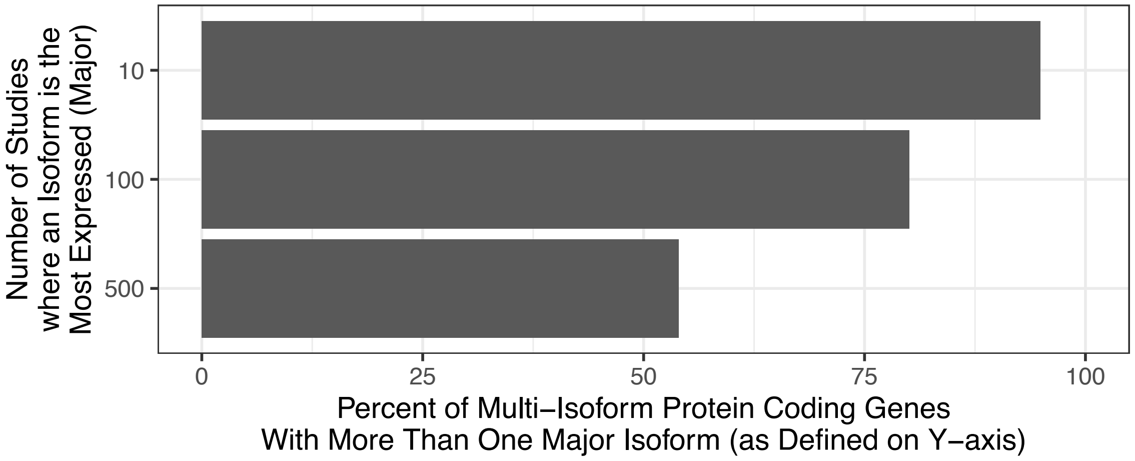


The percent of detected multi-isoform protein-coding genes with at least two major isoforms (x-axis) plotted against the minimum number of RNA-seq datasets where an isoform is the most expressed (10,291 human RNA-seq studies considered). Notably, for each RNA-seq dataset, we extracted the median isoform expression (TPM), making the estimate both conservative and robust. This shows that when RNA-seq is considered, the vast majority of genes utilize multiple isoforms. See Supplementary Table 6.

*Figure 2D.1*

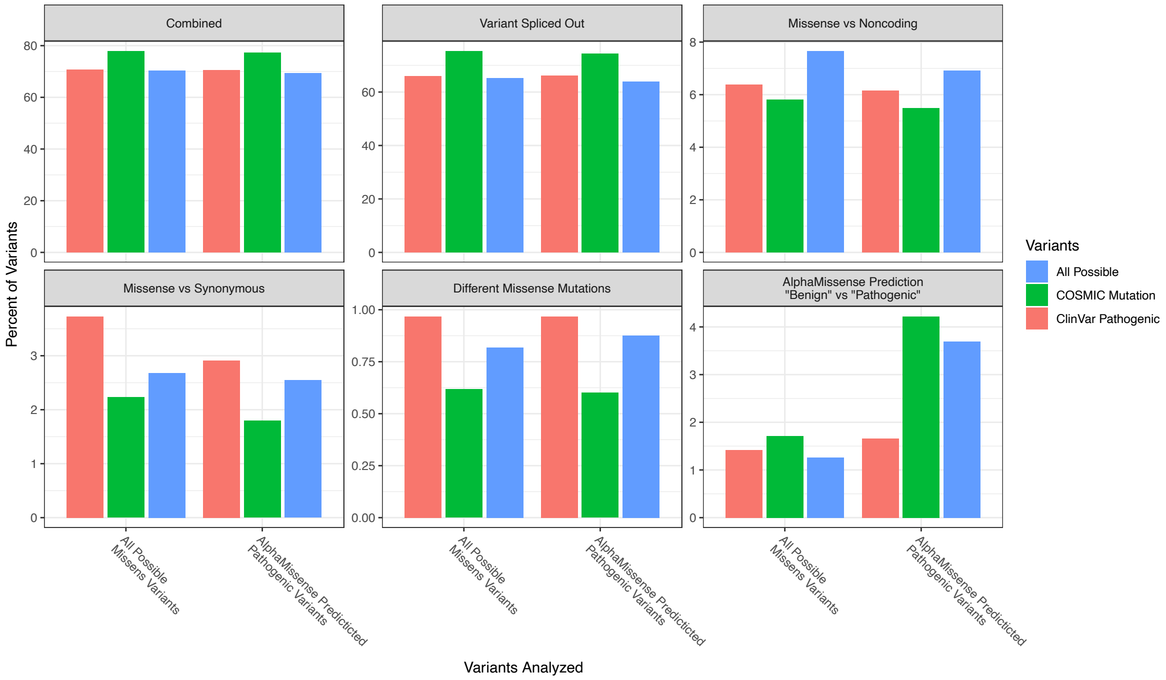


The frequency of isoform-dependent differences in variant interpretations (y-axis) for each category considered (subplots). The X-axis and color jointly indicate the subset of variants considered. Note that this is just a different visualization of the same data, as shown in the corresponding main figure.

*Figure 2D.2*

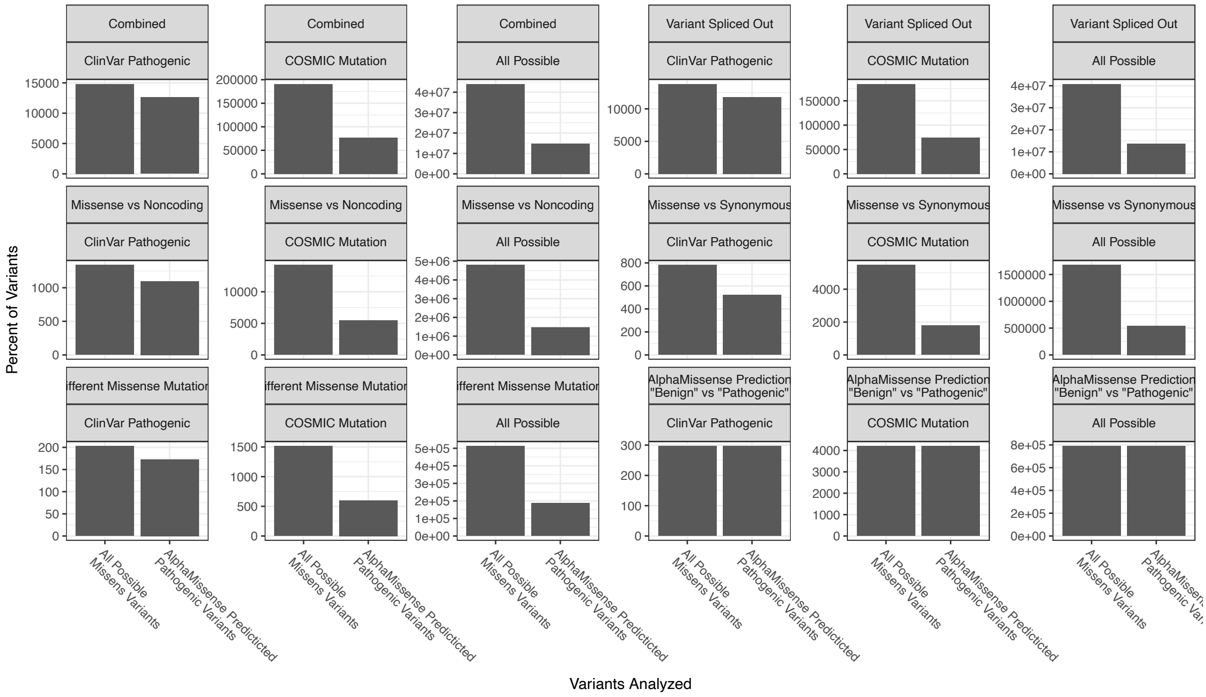

The number of variants with isoform-dependent differences in variant interpretations (y-axis) for each category. Subplots indicate the category (top grey bar) and the subset of variants considered (bottom grey bar). The subplot (indicated by the bottom grey bar) and the x-axis jointly indicate the subset of variants considered. This is the number of variants behind the fractions shown above and in the main figure.

*Figure 2D.3*

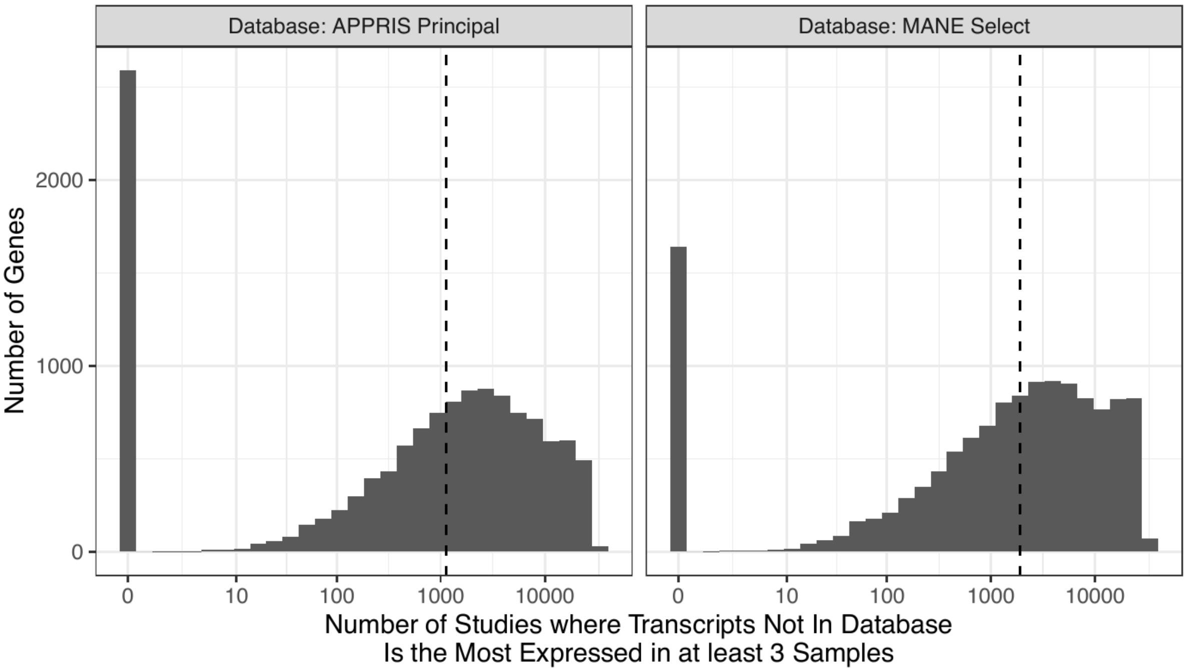


For each of the 10,291 human RNA-seq studies considered, we counted, for each gene, how many studies used transcripts not defined as "reference" by APPRIS or MANE Select (subplots) as the most expressed (major) isoform in at least 3 samples (x-axis). Only protein-coding genes found in all 3 datasets (n = 13,035) are shown. Dashed lines indicate the medians (1,139 and 1,897, respectively), and the x-axis is on a log10 scale. This shows that non-"reference" transcripts are used in many cells, tissues, and states and hence cannot be neglected. This analysis is a post-analysis of Figure 2A.7.

*Figure 2F.1*

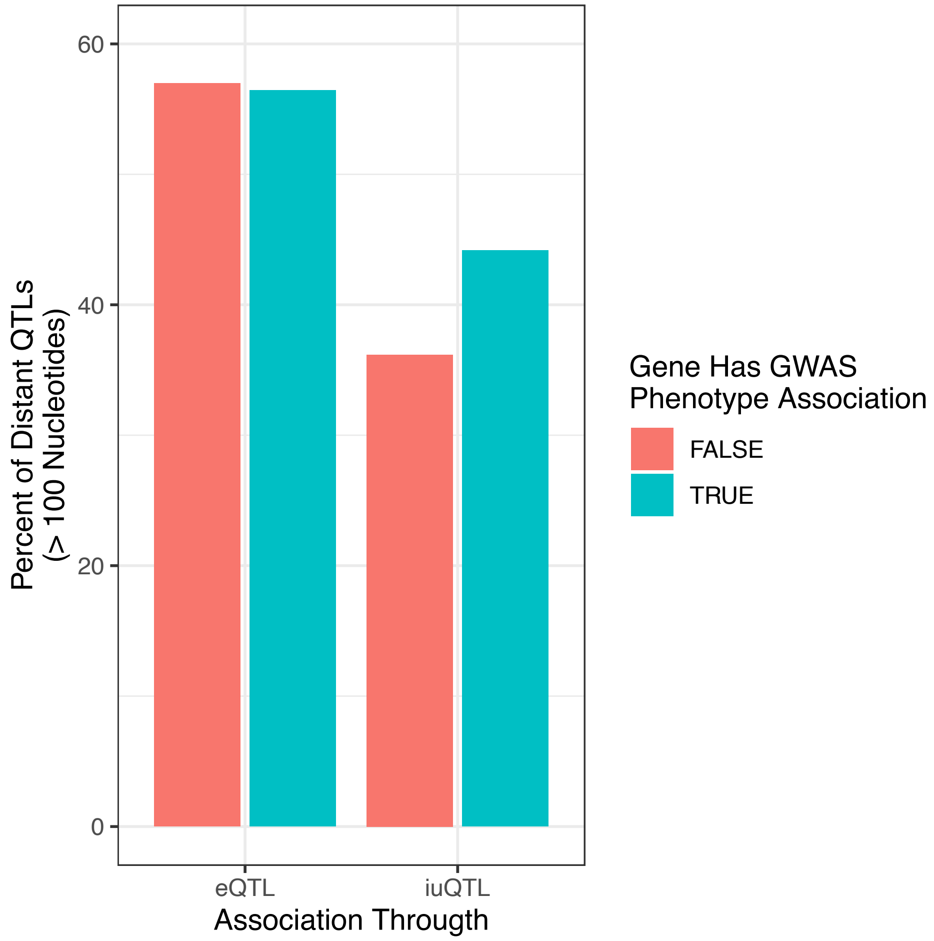


For eQTL and iuQTL (x-axis), the percent of QTLs that are more than 100 nucleotides away from the associated gene (distal, y-axis). The color indicates if the associated gene is also associated with a GWAS phenotype in our colocalization analysis. This shows that distal QTLs are similarly associated with GWAS phenotypes.
